# Unique neural signatures of childhood sexual abuse revisited

**DOI:** 10.1101/2025.02.10.25322009

**Authors:** Katharina Brosch, Vincent Hammes, Paula Usemann, Frederike Stein, Stephanie Zika, Franziska Schrott, Florian Thomas-Odenthal, Lea Teutenberg, Susanne Meinert, Katharina Thiel, Kira Flinkenflügel, Navid Schürmeyer, Janik Goltermann, Elisabeth J. Leehr, Linda M. Bonnekoh, Dominik Grotegerd, Nils Winter, Tim Hahn, Benjamin Straube, Hamidreza Jamalabadi, Andreas Jansen, Axel Krug, Udo Dannlowski, Igor Nenadić, Elvisha Dhamala, Tilo Kircher, Nina Alexander

## Abstract

Childhood sexual abuse (CSA) constitutes a detrimental subtype of childhood maltreatment (CM) associated with high trauma load and adverse health outcomes. Previous studies indicate CSA-specific reductions in gray matter volume (GMV) and cortical thickness. It remains challenging to disentangle brain alterations associated with CSA from those related to trauma intensity or psychopathology. Here, we apply a novel approach, comparing individuals with CSA, non-sexual maltreatment, and a non-maltreated control group, to identify CSA-specific findings.

Drawing from a cohort of *n*=2039 depressed and healthy men and women allowed us to match groups 1:1 for age, sex, depression diagnosis, and, for maltreated groups, trauma load. The Childhood Trauma Questionnaire was used to assess childhood maltreatment. Applying threshold-free cluster enhancement, we investigated GMV and cortical thickness in *n*=195 adults with CSA compared to non-sexually maltreated (*nCSA*, *n*=195) and non-maltreated (*nCM*, *n*=195) individuals.

*CSA* showed larger GMV in the right cerebellum compared to *nCSA* but not compared to *nCM*. *CSA* displayed larger cortical thickness encompassing the bilateral superior frontal gyri, pre- and postcentral gyri, supramarginal gyri, superior parietal cortices, precunei, and insulae, compared to both *nCSA* and *nCM*.

This is the largest study to investigate CSA-specific effects on brain morphometry, applying matched group comparisons. These findings highlight distinct neural signatures of CSA, characterized by preserved cortical thickness in regions also affected by major depression, and larger cerebellar GMV compared to non-sexual types of maltreatment. These results underscore the importance of distinguishing between types of childhood maltreatment and considering confounding factors when assessing their neurobiological impacts.

## Introduction

Childhood maltreatment (CM) is associated with long-term alterations in brain structure (1). CM is considered a heterogeneous entity of multiple adverse experiences during childhood, encompassing sexual, physical, and emotional abuse, and physical and emotional neglect (2). These subtypes vary in degree of threat and deprivation (3,4). Threat constitutes harm to one’s bodily integrity, whereas deprivation constitutes the absence of adequate environmental stimulation (5). Childhood sexual abuse (CSA) is a particularly detrimental threat subtype of CM and associated with adverse and long-lasting mental and physical health outcomes (6). Worldwide, one in 5 women and one in 13 men report experiences of CSA (7).

Survivors of CSA have an increased risk of developing several psychiatric disorders, such as major depressive disorder (MDD), anxiety disorder, post-traumatic stress disorder (PTSD), borderline personality disorder, schizophrenia, eating disorders, conversion disorder, and somatoform disorders (6). Adverse physical health outcomes of CSA include increased risk for fibromyalgia and obesity (6,8–10).

While most CM research has focused on investigating the general adverse effects of all different subtypes combined, previous studies have also demonstrated differential effects of specific trauma subtypes on outcome domains such as structural and functional brain changes, inflammation, and emotion dynamics (4,11–14). Identifying such differential outcomes may inform targeted prevention and intervention strategies to optimally treat adverse outcomes of specific CM subtypes (15).

Neuroimaging approaches have been central in CM research: during sensitive phases of brain development in childhood and adolescence, environmental influences can be particularly impactful (4). A recent meta-analysis summarizing 45 studies found consistent evidence of smaller cortical thickness and gray matter volume (GMV) in the median cingulate and paracingulate gyri, associated with general CM experiences. Further, smaller GMV in the supplementary motor area and smaller cortical thickness in the anterior cingulate/paracingulate gyri and left middle frontal gyrus were reported in individuals with CM (1).

However, structural CM findings remain inconsistent across individual studies, which has been explained by factors such as heterogeneous cohort demographics, timing of CM, frequency of exposure, and specific subtype of CM (1,4,11,16,17). In CSA, MRI studies that included psychiatric patients and healthy controls reported unique morphometric signatures (18). Specifically, earlier studies identified brain morphometric alterations in CSA survivors compared to non-victimized controls in the hippocampus, the frontal cortex, the dorsal anterior cingulate cortex, and the corpus callosum (19–23). Importantly, the group differences in brain structure identified in studies comparing CSA to no CM may reflect neural correlates of general maltreatment exposure rather than specific effects of CSA (24,25). These neural differences could potentially be observed across different types of maltreatment.

The few studies exploring the *specificity* of CSA by explicitly comparing the effects of different types of CM have yielded inconsistent results. Investigating cortical thickness, Heim et al. examined the differential effects of CM subtypes in a sample of adult women (26). Here, CSA was associated with cortical thinning in the genital representation field of the primary somatosensory cortex when controlling for other types of CM and depression using regression analysis (26). Tomoda et al. (27) demonstrated reduced GMV in the visual cortex of young women exposed to CSA compared to healthy controls without CSA exposure. Respective effects were found to be partly stable when excluding CSA individuals with psychopathology. Other studies further reported significantly smaller cortical thickness in the left rostral middle frontal gyrus, the bilateral fusiform gyri, and the right supramarginal gyrus and reduced GMV in the middle occipital gyrus in individuals exposed vs. not exposed to CSA (25,28). Concurrently, these effects were not observed for other types of CM. In patients with MDD, CSA and other types of CM were negatively correlated with entorhinal thickness and hippocampal volume (29).

Previous literature on CSA-specific effects on brain morphology has generally relied on smaller sample sizes (with less than *n*=35 or unreported numbers of individuals exposed to CSA in individual studies) that may not generalize across different populations (26,27). Additional considerations impede the interpretability of CSA-specific effects in current research, which are 1) dose-dependent effects (i.e. confounding effects of higher trauma load) and 2) effects of psychopathology.

First, some studies suggest that CSA is related to more negative health outcomes than other types of CM (30). However, this finding might not be a consequence of the specific exposure to CSA *per se* but could result from a generally higher trauma load observed in CSA survivors (30). Dose-dependent effects of CM have been reported before, and CM *severity* was associated with more severe psychopathological symptoms and increased cortical thinning (9,31–33). Therefore, studies should account for general trauma load when investigating CSA-specific effects.

Second, it remains challenging to disentangle structural brain changes associated with CSA from those associated with psychopathology. Studies including CSA survivors who meet the criteria for specific psychiatric disorders may not provide unbiased findings on CSA effects per se (24,27). Conversely, as CSA is associated with a significant risk for psychiatric illness, such as MDD, including only psychiatrically healthy survivors of CSA would introduce a bias. Findings from healthy, maltreated individuals might underestimate the effects of CSA and confound findings with brain alterations associated with resilience to CM (27,34). Studies investigating specific CSA effects should therefore include CSA survivors with and without a history of psychiatric illnesses and account for diagnosis effects.

In sum, we have an insufficient understanding of the specific effects of CSA on brain morphometry, as previous studies are primarily based on small sample sizes, and specific methodological considerations remain unaddressed. Previous work has repeatedly called for the replication of CSA-specific brain imaging studies and the consideration of trauma load (4,6).

Here, we utilize the data from the large German FOR2107 consortium, which investigates the neurobiology of major psychiatric disorders (35). This trauma-enriched cohort allows for the granular investigation of CSA effects. Using this deeply phenotyped sample, we are uniquely positioned to implement a study design that can compare individuals with CSA to a) non-sexually maltreated individuals and b) non-maltreated individuals. Our groups are carefully matched 1:1 regarding trauma load (in maltreated groups), age, diagnostic group (healthy and depressed), and sex distribution to identify unique effects of CSA on cortical thickness and GMV.

## Methods and Material

### Sample

*N*=585 healthy and depressed participants from a total cohort of *n* = 2039 were selected for the current study. We compared three groups of participants, matched 1:1, each *n* = 195: 1) *CSA*, 2) other forms of CM but not CSA (*nCSA*), and 3) no experience of CM (*nCM*). The three groups were age, sex, and MDD diagnosis-matched to the CSA group. Participants were drawn from the Marburg-Münster Affective Disorder Cohort Study (MACS), a bi-center cohort study and part of the FOR2107 consortium investigating the neurobiology of major psychiatric disorders (35).

Participants were White, with Central-European ancestry, aged 18-65, and were recruited via newspapers, advertisements, and flyers. Patients were additionally recruited from in- and out-patient departments of the German universities of Marburg and Münster, as well as local German psychiatric hospitals. Participants were phenotyped using a comprehensive questionnaire battery, neuropsychological assessment, neuroimaging, and collection of biomaterials (see (35) for a detailed description). Trained staff applied the German version of the structured clinical interview (SCID-I) for the DSM-IV-TR (36). Exclusion criteria included substance dependence, neurological or other severe somatic disorders, traumatic head injury, a verbal IQ ≤ 80 (assessed using the multiple-choice vocabulary test, MWT-B (37)), and MRI contraindications (e.g., pregnancy, ferromagnetic implants, claustrophobia). Individuals with and without a history of MDD were included. Individuals classified as psychiatrically healthy did not meet the criteria for any current or past psychiatric disorders according to DSM-IV-TR. Participants gave written, informed consent and received financial compensation. The study protocols were approved by the local Ethic Committees in Marburg and Münster, Germany, per the declaration of Helsinki.

### Assessment of CM

Maltreatment experienced during childhood was assessed retrospectively using the German version of the Childhood Trauma Questionnaire (CTQ) (38). This self-report questionnaire assesses CM across five subscales (physical neglect, emotional neglect, physical abuse, emotional abuse, and sexual abuse) with 25 items, 5 per subscale, summed up to a CM sum score, ranging from 25 - 125. The CTQ further includes three additional items assessing minimalization and denial. Items are rated on a 5-point Likert scale, with higher scores indicating more severe or frequent maltreatment. Individuals were grouped in the *CSA* group when the validated cut-off score for the sexual abuse subscale was met (sexual abuse ≥ 8) (30). Sexual abuse scale items included both touch and non-touch experiences of sexual abuse. When individuals met any other maltreatment subscale cut-off score (physical abuse ≥ 8, emotional abuse ≥ 10, physical neglect ≥ 8, emotional neglect ≥ 15), while the CSA criterion was not met, individuals were classified as *nCSA* (non-sexual forms of maltreatment) (30). If none of the cut-off scores were met, individuals were classified as *nCM* (no form of CM).

### Sample selection and matching

In the cohort of *n* = 2039, *n* = 204 met the criteria for CSA, while *n* = 795 were classified as non-sexually maltreated (*nCSA*), and *n* = 1040 were classified as non-maltreated (*nCM*) according to cut-off scores. We then matched the *nCSA* and *nCM* group 1:1 to the *CSA* group regarding age, sex, diagnostic group, and, in the case of *nCSA*, general trauma load (CTQ sum score).

Matching for trauma load proved difficult, as individuals with *CSA* (*M* = 59.28, *SD* = 18.48) reported significantly higher CTQ sum scores compared to the *nCSA* group (*M* = 45.45, *SD* = 11.03). As a result, we were unable to match non-sexually abused individuals to the entire CSA group (*n*=204), albeit drawing from a sample of *n*=795 non-sexually abused individuals. This finding is in line with previous observations that CSA is associated with higher general trauma load (30).

To achieve successful matching, one of our main objectives, we excluded *n*=9 *CSA* individuals who reported the most severe CM (CTQ sum scores > 93, with 125 being the maximum). Our final sample included *N* = 585 individuals with *n* = 195 each in the *CSA*, *nCSA,* and *nCM* groups; see Figure 1 for the three groups and applied comparisons, and Figure 2 below for sample selection.

**Figure 1:**
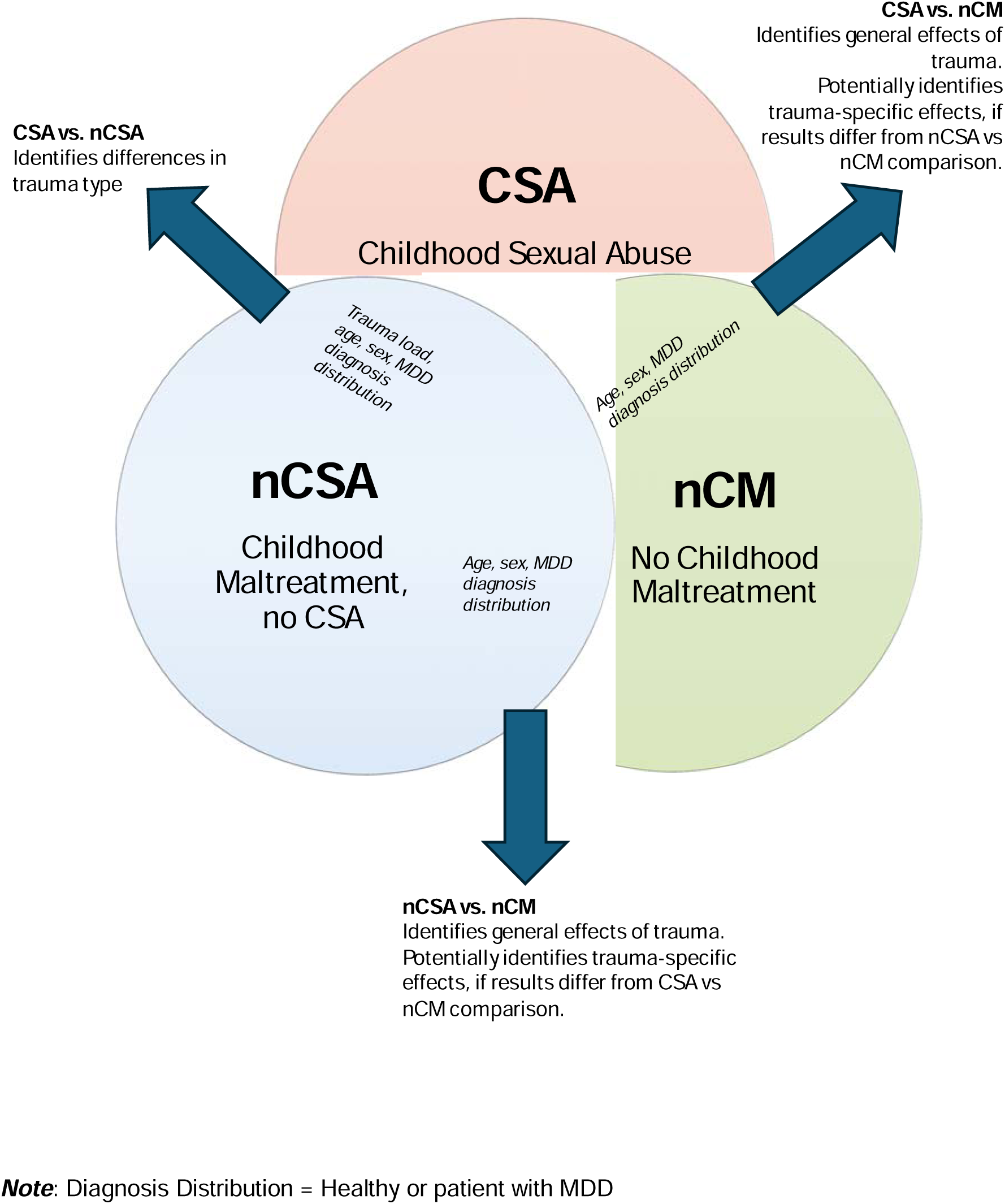
Visualization of the three groups, possible comparison outcomes and interpretation of possible group comparisons.

**Figure 2:**
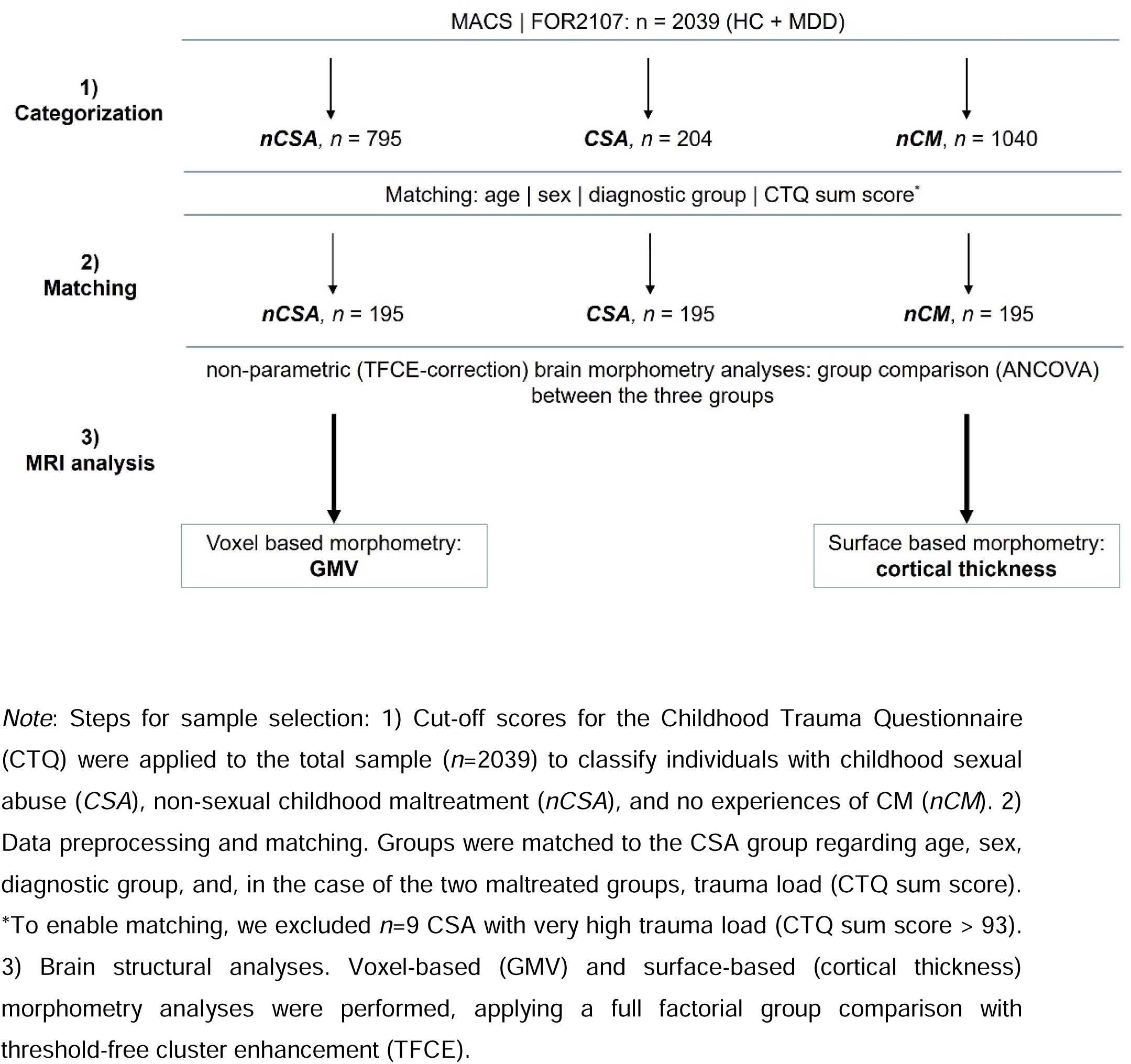
Sample selection and analysis pipeline.

The final CSA sample (*n =* 195) was comprised of mostly women (86%) and patients with MDD (77%), with a mean age of 41.5 years; a naturalistic representation of the actual distribution of CSA in men and women and its effects on mental health (6,7).

### Group comparisons

The goal of this study was to identify unique neural correlates of CSA, while accounting for confounding factors such as trauma load, depression diagnosis, age, and sex. To this end, we chose a 1×3 design with three matched groups: *CSA*, *nCSA*, and *nCM*. Comparisons between the three groups enables a comprehensive understanding of CSA-specific neural correlates, i.e., qualitative maltreatment subtype differences (*CSA* vs. *nCSA*), broader impact of CSA on the brain (*CSA* vs. *nCM*), and general maltreatment effects (*nCSA* vs *nCM*) in individuals with and without a history of depression. Contrasting findings from these three group comparisons allow for a deeper understanding which brain alterations are shared or unique in CSA and other types of CM (Figure 1). To avoid confounding effects of MDD, the *nCM* group does not constitute a “traditional” healthy control group, as this group was also matched to *CSA* group characteristics and includes mostly women and individuals with a history of MDD. Our framework ensures that findings are not misattributed to general trauma load, psychopathology, or shared maltreatment effects, making it uniquely powerful to disentangle the complex relationships between maltreatment types, trauma load, and neurodevelopmental outcomes.

### MRI data acquisition and preprocessing

In Marburg and Münster, Germany, high-resolution T1 images were acquired with a 3T whole-body MRI scanner (Marburg: 12-channel head matrix Rx-coil, Tim Trio, Siemens, Erlangen, Germany; Münster: 20-channel head matrix Rx-coil, Prisma, Siemens, Erlangen, Germany) (39). Parameters of the 3D-fast gradient echo sequence (MPRAGE) varied between sites. Marburg: field of view (FOV) = 256 mm, 176 sagittal slices, repetition time (TR) = 1900 ms, echo time (TE) = 2.26 ms, inversion time (TI) = 900 ms, slice thickness = 1 mm, voxel size = 1 × 1 × 1 mm, flip angle = 9°, Münster: FOV = 256 mm, 192 sagittal slices, TR = 2130 ms, TE = 2.28 ms, TI = 900 ms, slice thickness = 1 mm, voxel size = 1 × 1 × 1 mm, flip angle 8°.

Preprocessing was performed by applying default parameters of the CAT12 toolbox (build 1184, Gaser, Structural Brain Mapping Group, Jena University Hospital, Jena, Germany) implemented in SPM12 (v7771, Statistical Parametric Mapping, Institute of Neurology, London, UK) running under MATLAB (version v2017a, The MathWorks, USA). Images passed individual quality control, with a senior researcher manually checking for artifacts, abnormalities, and image quality. Brain images were segmented into gray matter, white matter, and cerebrospinal fluid and then normalized using an adopted DARTEL algorithm (40). For data smoothing, 8-mm (for GMV) and 20-mm (for cortical thickness) full-width half-maximum Gaussian kernels with an absolute threshold of 0.1 were applied.

### Statistical analyses

Nearest Neighbor matching (using the MatchIt package 4.5.0) was performed to match group sizes 1:1 to the smallest group (*CSA*) regarding age, sex, diagnostic group, and, in the case of the *nCSA* group, CTQ sum score (41). All non-brain statistical analyses were performed using R (R Core Team (2021), version 4.1.2). To assess group differences in demographic data, analysis of variance (ANOVA) with subsequent post-hoc Tukey Honest Significant Differences (TukeyHSD) tests were conducted for continuous variables and Pearson’s chi-squared test with subsequent post-hoc analyses based on the residuals were computed for categorical variables (see Table 1). To further evaluate differences and comparability between the groups, Bayesian factor analysis was conducted to quantify evidence for the null and alternative hypothesis (see Table 1 and Supplementary Table 1). Here, Bayesian ANOVA was performed to evaluate evidence for group effects in continuous variables, and Bayesian contingency table analysis was applied for categorical variables.

**Table 1.**
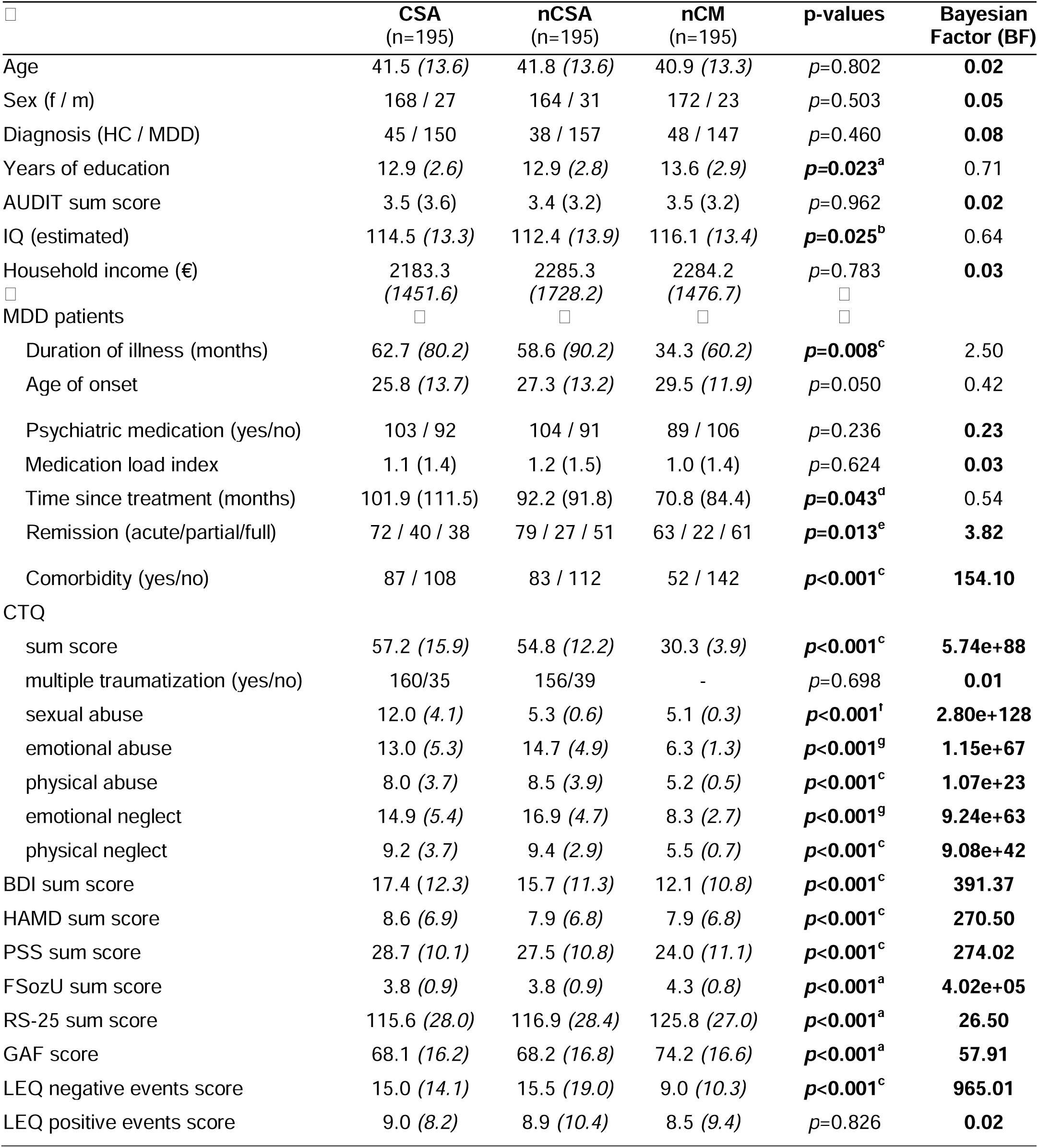
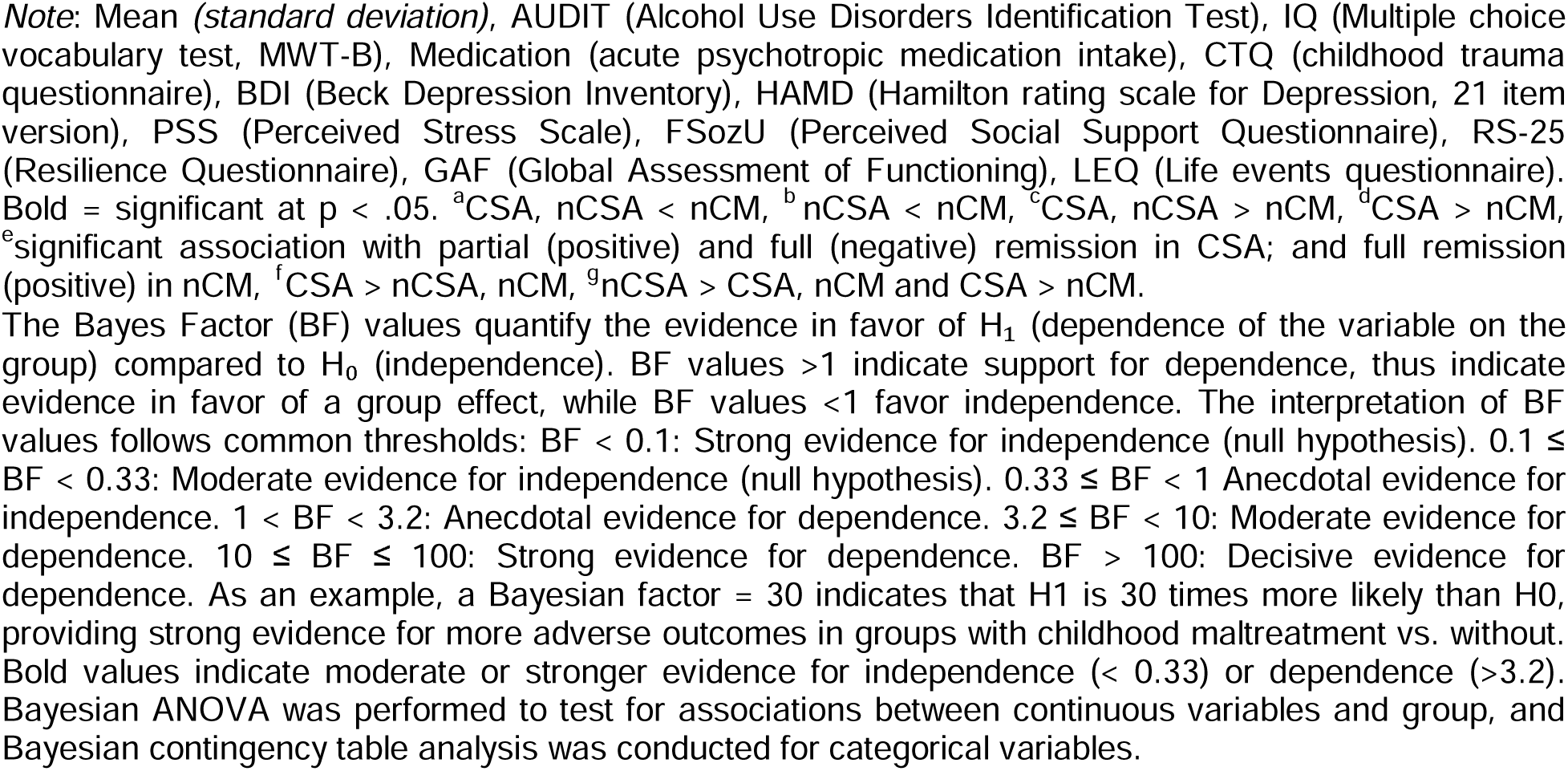
Sample Characteristics (N=585)

| □ | CSA<br>(n=195) | nCSA<br>(n=195) | nCM<br>(n=195) | p-values | Bayesian<br>Factor (BF) |
| --- | --- | --- | --- | --- | --- |
| Age | 41.5 (13.6) | 41.8 (13.6) | 40.9 (13.3) | $p=0.802$ | **0.02** |
| Sex (f / m) | 168 / 27 | 164 / 31 | 172 / 23 | $p=0.503$ | **0.05** |
| Diagnosis (HC / MDD) | 45 / 150 | 38 / 157 | 48 / 147 | $p=0.460$ | **0.08** |
| Years of education | 12.9 (2.6) | 12.9 (2.8) | 13.6 (2.9) | **$p=0.023^a$** | 0.71 |
| AUDIT sum score | 3.5 (3.6) | 3.4 (3.2) | 3.5 (3.2) | $p=0.962$ | **0.02** |
| IQ (estimated) | 114.5 (13.3) | 112.4 (13.9) | 116.1 (13.4) | **$p=0.025^b$** | 0.64 |
| Household income (€) | 2183.3<br>(1451.6) | 2285.3<br>(1728.2) | 2284.2<br>(1476.7) | $p=0.783$<br>□ | **0.03**<br>□ |
| □<br>MDD patients | □ | □ | □ | □ |  |
| Duration of illness (months) | 62.7 (80.2) | 58.6 (90.2) | 34.3 (60.2) | **$p=0.008^c$** | 2.50 |
| Age of onset | 25.8 (13.7) | 27.3 (13.2) | 29.5 (11.9) | $p=0.050$ | 0.42 |
| Psychiatric medication (yes/no) | 103 / 92 | 104 / 91 | 89 / 106 | $p=0.236$ | **0.23** |
| Medication load index | 1.1 (1.4) | 1.2 (1.5) | 1.0 (1.4) | $p=0.624$ | **0.03** |
| Time since treatment (months) | 101.9 (111.5) | 92.2 (91.8) | 70.8 (84.4) | **$p=0.043^d$** | 0.54 |
| Remission (acute/partial/full) | 72 / 40 / 38 | 79 / 27 / 51 | 63 / 22 / 61 | **$p=0.013^e$** | **3.82** |
| Comorbidity (yes/no) | 87 / 108 | 83 / 112 | 52 / 142 | **$p<0.001^c$** | **154.10** |
| CTQ |  |  |  |  |  |
| sum score | 57.2 (15.9) | 54.8 (12.2) | 30.3 (3.9) | **$p<0.001^c$** | **5.74e+88** |
| multiple traumatization (yes/no) | 160/35 | 156/39 | - | $p=0.698$ | **0.01** |
| sexual abuse | 12.0 (4.1) | 5.3 (0.6) | 5.1 (0.3) | **$p<0.001^f$** | **2.80e+128** |
| emotional abuse | 13.0 (5.3) | 14.7 (4.9) | 6.3 (1.3) | **$p<0.001^g$** | **1.15e+67** |
| physical abuse | 8.0 (3.7) | 8.5 (3.9) | 5.2 (0.5) | **$p<0.001^c$** | **1.07e+23** |
| emotional neglect | 14.9 (5.4) | 16.9 (4.7) | 8.3 (2.7) | **$p<0.001^g$** | **9.24e+63** |
| physical neglect | 9.2 (3.7) | 9.4 (2.9) | 5.5 (0.7) | **$p<0.001^c$** | **9.08e+42** |
| BDI sum score | 17.4 (12.3) | 15.7 (11.3) | 12.1 (10.8) | **$p<0.001^c$** | **391.37** |
| HAMD sum score | 8.6 (6.9) | 7.9 (6.8) | 7.9 (6.8) | **$p<0.001^c$** | **270.50** |
| PSS sum score | 28.7 (10.1) | 27.5 (10.8) | 24.0 (11.1) | **$p<0.001^c$** | **274.02** |
| FSozU sum score | 3.8 (0.9) | 3.8 (0.9) | 4.3 (0.8) | **$p<0.001^a$** | **4.02e+05** |
| RS-25 sum score | 115.6 (28.0) | 116.9 (28.4) | 125.8 (27.0) | **$p<0.001^a$** | **26.50** |
| GAF score | 68.1 (16.2) | 68.2 (16.8) | 74.2 (16.6) | **$p<0.001^a$** | **57.91** |
| LEQ negative events score | 15.0 (14.1) | 15.5 (19.0) | 9.0 (10.3) | **$p<0.001^c$** | **965.01** |
| LEQ positive events score | 9.0 (8.2) | 8.9 (10.4) | 8.5 (9.4) | $p=0.826$ | **0.02** |

Note: Mean (standard deviation), AUDIT (Alcohol Use Disorders Identification Test), IQ (Multiple choice vocabulary test, MWT-B), Medication (acute psychotropic medication intake), CTQ (childhood trauma questionnaire), BDI (Beck Depression Inventory), HAMD (Hamilton rating scale for Depression, 21 item version), PSS (Perceived Stress Scale), FSozU (Perceived Social Support Questionnaire), RS-25 (Resilience Questionnaire), GAF (Global Assessment of Functioning), LEQ (Life events questionnaire). Bold = significant at $p < .05$ . <sup>a</sup>CSA, nCSA < nCM, <sup>b</sup>nCSA < nCM, <sup>c</sup>CSA, nCSA > nCM, <sup>d</sup>CSA > nCM, <sup>e</sup>significant association with partial (positive) and full (negative) remission in CSA; and full remission (positive) in nCM, <sup>f</sup>CSA > nCSA, nCM, <sup>g</sup>nCSA > CSA, nCM and CSA > nCM.
The Bayes Factor (BF) values quantify the evidence in favor of $H_1$ (dependence of the variable on the group) compared to $H_0$ (independence). BF values >1 indicate support for dependence, thus indicate evidence in favor of a group effect, while BF values <1 favor independence. The interpretation of BF values follows common thresholds: BF < 0.1: Strong evidence for independence (null hypothesis). $0.1 \leq \text{BF} < 0.33$ : Moderate evidence for independence (null hypothesis). $0.33 \leq \text{BF} < 1$ : Anecdotal evidence for independence. $1 < \text{BF} < 3.2$ : Anecdotal evidence for dependence. $3.2 \leq \text{BF} < 10$ : Moderate evidence for dependence. $10 \leq \text{BF} \leq 100$ : Strong evidence for dependence. BF > 100: Decisive evidence for dependence. As an example, a Bayesian factor = 30 indicates that $H_1$ is 30 times more likely than $H_0$ , providing strong evidence for more adverse outcomes in groups with childhood maltreatment vs. without. Bold values indicate moderate or stronger evidence for independence (< 0.33) or dependence (>3.2). Bayesian ANOVA was performed to test for associations between continuous variables and group, and Bayesian contingency table analysis was conducted for categorical variables.

### Brain structural analyses

We compared GMV and cortical thickness in the three groups (*CSA*, *nCSA*, *nCM*). For the voxel-based morphometry (VBM) analyses investigating GMV, we implemented whole-brain analysis of covariance (ANCOVA) models using a full-factorial design in SPM, including the covariates age, sex, diagnosis, total intracranial volume (TIV) and, per our MRI quality assurance protocol, gradient coil and body coil changes in Marburg, which also account for site effects (39). For the surface-based morphometry (SBM) analyses investigating cortical thickness, we used the CAT12 toolbox (CAT12.8.2, version r2159) that builds on SPM with the same basic model and covariates as in the VBM analyses, without TIV as a covariate. For both modalities, we used threshold-free cluster enhancement (TFCE), a non-parametric multiple-comparison correction, using the TFCE Toolbox (version r264) (42). We applied the Smith Method with 20,000 permutations and subsequent family-wise error (FWE) correction for multiple comparisons. Results were considered significant at α < 0.05. Anatomical labeling of clusters was performed using the Dartel space Neuromorphometrics atlas implemented in SPM12 for voxel-based morphometry analyses and the Desikan-Killiany DK40 atlas for surface-based morphometry analyses (43). Although TFCE does not allow for the extraction of effect sizes, we converted TFCE values to t-values, which we then converted to Cohen’s d effect sizes to obtain approximate effect sizes of significant effect. We note that this is an approximation, and these values only represent the peak-voxel, and actual effects might be smaller.

## Results

The final sample included 585 participants (mean age = 41.4, *SD* = 13.49 years), with 195 participants per group (*CSA, nCSA, and nCM*). Clinical and demographic characteristics are shown below in Table 1. Matching was successful, as indicated by no significant differences in the groups in age, sex, and diagnosis distribution. *CSA* and *nCSA* did not significantly differ in their CTQ sum scores. Compared to the CSA group, the *nCSA* group was characterized only by significantly lower sexual abuse and higher emotional neglect and abuse (Supplementary Table 1).

As shown in the sample characteristics below, the behavioral outcomes demonstrated a pervasive, adverse effect of CM that is sustained even when diagnosis is accounted for. The *CSA* and the *nCSA* groups show significantly higher self-reported and rater-based depressive symptoms, higher perceived stress, higher experiences of negative stressful life events, lower social support, lower resilience, fewer years of education, and lower global functioning compared to the *nCM* group (see Table 1). This is further supported by large Bayesian factor scores (>10) in multiple adverse outcomes. These findings align with current literature (44,45).

### GMV analyses

To identify CSA specific GMV alterations, we first compared the *CSA* to the *nCSA* group. Second, we investigate results of the *CSA* vs. *nCM* comparison and contextualize findings using the *nCSA* vs. *nCM* group comparison. Covariates in all models include age, sex, diagnosis, site, and TIV. We first describe findings for positive contrasts for the three comparisons, and then the negative contrasts. We describe clusters with *k* > 100 voxels here, a full list of all significant results can be found in the Supplement.

#### CSA > nCSA

The *CSA* group exhibited significantly larger GMV in the right exterior cerebellum (*k*=1225, TFCE=917.92, *p*_FWE_=0.025, x/y/z=33/-48/-32) compared to the *nCSA* group (Figure 3A, Supplementary Table 2. The approximate effect size for this effect (converted from t-statistic), is *d*=0.38, indicating a small effect. These findings may signify brain alterations due to qualitative maltreatment experiences. To further describe respective findings, we explored eigenvariate values for all three groups (Supplementary Figure 1). Significant differences cluster emerged only in the *CSA* > *nCSA* comparison, whereas *nCSA* and *CSA* do not differ significantly in GMV compared to *nCM* in the respective comparisons. These findings indicate nuanced differences in the right cerebellum dependent on maltreatment subtype.

**Figure 3:**
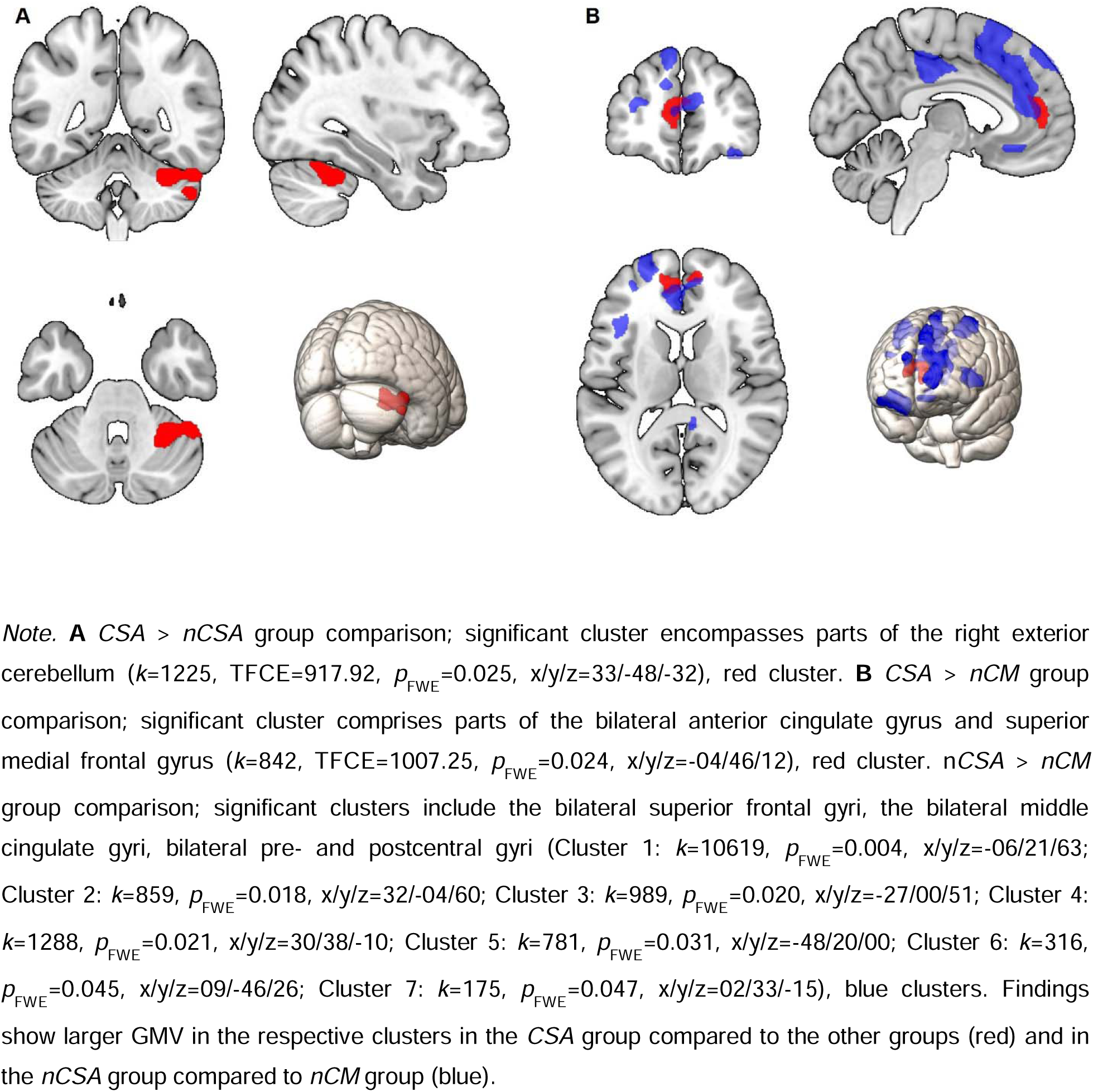
Clusters exhibiting significantly larger GMV (TFCE, *p*_FWE_-corrected) in **A** *CSA*.

#### CSA > nCM

Compared to the *nCM* group, the *CSA* group exhibited larger GMV in the bilateral anterior cingulate gyrus and bilateral superior medial frontal gyrus (mSFG) (*k*=842, TFCE=1007.25, *p*_FWE_=0.024, x/y/z=-04/46/12, Figure 3B, Supplementary Table 3). The approximate effect size for this effect (converted from t-statistic), is *d*=0.41, indicating a small effect. Similar regions were detected for the *nCSA* > *nCM* comparison, indicating possible general maltreatment effects when comparing individuals with and without MDD.

#### nCSA > nCM

The *nCSA* group exhibited larger GMV in seven large clusters, including the bilateral superior frontal gyri, the bilateral middle cingulate gyri, bilateral pre- and postcentral gyri (Figure 3B, Supplementary Table 4). Cluster 1: *k*=10619, TFCE=1386.31, *p*_FWE_=0.004, x/y/z=-06/21/63; Cluster 2: *k*=859, TFCE=1067.21, *p*_FWE_=0.018, x/y/z=32/-04/60; Cluster 3: *k*=989, TFCE=1048.91, *p*_FWE_=0.020, x/y/z=-27/00/51; Cluster 4: *k*=1288, TFCE=1041.91, *p*_FWE_=0.021, x/y/z=30/38/-10; Cluster 5: *k*=781, TFCE=957.70, *p*_FWE_=0.031, x/y/z=-48/20/00; Cluster 6: *k*=316, TFCE=883.27, *p*_FWE_=0.045, x/y/z=09/-46/26; Cluster 7: k=175, TFCE=874.00, *p*_FWE_=0.047, x/y/z=02/33/-15. These findings may indicate brain changes in non-sexual forms of CM in a population of individuals with and without MDD.

#### CSA < nCM; CSA < nCSA; nCSA < nCM

We did not detect any significant clusters for these comparisons, p > 0.05.

To rule out possible underlying sex or diagnosis effects interacting with the groups and driving our findings, we ran sex and MDD diagnosis (dichotomous HC vs. MDD) interaction analyses. These analyses revealed no significant interaction effects on GMV. The results suggest that neither MDD diagnosis nor sex impact the significant findings.

Moreover, we investigated the potential influence of comorbidity on the observed differences in GMV. To this end, we repeated the analyses, including comorbidity (dichotomous, no vs. yes) as an additional covariate, yielding similar results. This indicates that comorbidity does not account for the observed effects (Supplementary Figure 2 with Supplementary Table 5 - 7).

### Cortical thickness analyses

Analogously to GMV, we compared the *CSA* to the *nCSA* group to identify specific alterations in cortical thickness related to CSA. Next, we investigated results of the *CSA* vs. *nCM*, and contextualized findings using the *nCSA* vs. *nCM* comparison (see Supplement). Covariates in all models include age, sex, site, and diagnosis. We first describe findings for positive contrasts for the three comparisons, and then the negative contrasts. We describe clusters with *k* > 100 voxels here, a full list of all significant results can be found in the Supplement.

#### CSA > nCSA

We identified three clusters of larger cortical thickness in the *CSA* group compared to the *nCSA* group, encompassing regions of the bilateral superiorfrontal gyrus, pre- and postcentral gyri, supramarginal gyrus, superior parietal cortex, precuneus and insula (Cluster 1: *k*=11517, TFCE=19182.83, *p*_FWE_=0.011, x/y/z=31/20/08; Cluster 2: *k*=13137, TFCE=16037.30, *p*_FWE_=0.017, x/y/z=-21/-83/39; Cluster 3: *k*=246, TFCE=9500.77, *p*_FWE_=0.049, x/y/z=06/44/26) (Figure 4A). Supplementary Table 8 lists all clusters and their respective atlas region. Approximated Cohen’s d effect sizes were Cluster 1: *d*=0.39, Cluster 2: *d*=0.37, and Cluster 3: *d*=0.22, all indicating small effects. These findings may signify brain alterations due to qualitative maltreatment experiences.

**Figure 4:**
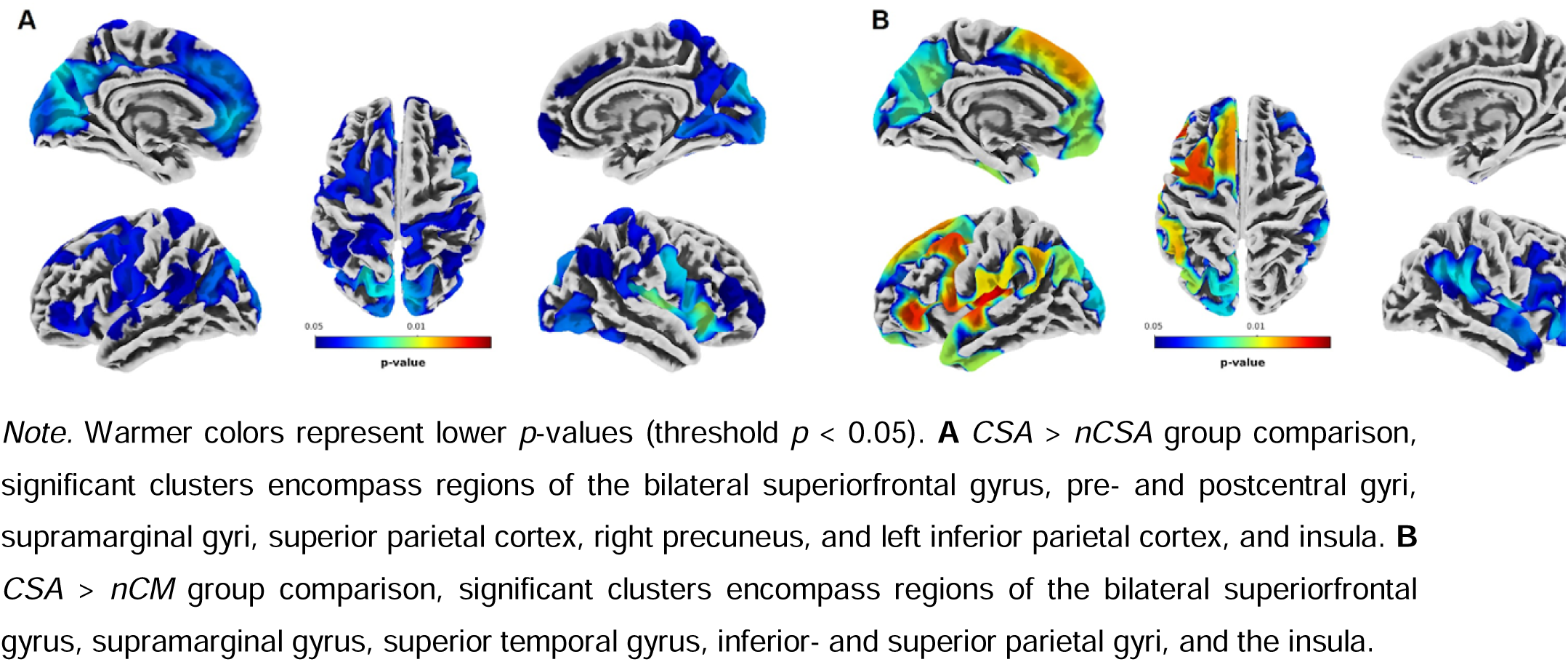
Clusters exhibiting significantly larger cortical thickness (TFCE, p_FWE_- corrected) in *CSA* compared *to **A** nCSA and **B** nCM*.

#### CSA > nCM

The *CSA* group exhibited greater cortical thickness compared to the *nCM* group in two clusters encompassing bilateral superiorfrontal, supramarginal, superior temporal, and inferior parietal regions in addition to the insula (Cluster 1: *k*=13400, TFCE=27566.17, *p*_FWE_=0.005, x/y/z=-29/04/54; Cluster 2: *k*=6478, TFCE=18517.88, *p*_FWE_ =0.018, x/y/z=63/-40/24) (Figure 4B). Supplementary Table 9 lists all clusters and their respective atlas region. The approximated Cohen’s d effect sizes were Cluster 1: *d*=0.37, and Cluster 2: *d*=0.35, indicating small effect sizes.

#### nCSA > nCM

There was no significant finding for cortical thickness for this comparison, p > 0.05.

#### CSA < nCSA; CSA < nCM; nCSA < nCM

There were no significant findings for cortical thickness for these comparisons, p > 0.05.

Significant clusters in the *CSA* > *nCSA* and *CSA* > *nCM* comparisons largely overlapped, indicating possible unique cortical thickness effects that distinguish CSA from both *nCSA* and *nCM*. The identified clusters in cortical thickness have been previously associated with major depression (46). To further explore these findings, we conducted additional exploratory analyses, including the Hamilton rating scale for Depression (HAMD) score as a covariate to investigate if depression *severity* impacted our findings. When accounting for depression severity, significant differences in cluster sizes between *CSA* vs. *nCSA* diminished, whereas differences between *CSA* and *nCM* became more pronounced (Supplementary Figure 3, Supplementary Table 10 and 11). This points to an interaction effect of depression severity and maltreatment type for cortical thickness.

As with GMV, we investigated possible interaction effects of sex or MDD diagnosis (dichotomous HC vs. MDD) by maltreatment group. Interaction analyses revealed no significant effects. Furthermore, we investigated the potential influence of comorbidity on the observed differences in cortical thickness. To this end, we included comorbidity (dichotomous, no vs. yes) as an additional covariate in the model, which yielded similar results, indicating that comorbidity does not influence the observed effects (Supplementary Figure 4, Supplementary Table 12 and 13).

In summary, our findings suggest that CSA is characterized by larger cortical thickness in frontoparietal areas and larger cerebellar GMV when compared to individuals with non-sexual forms of maltreatment.

## Discussion

This is the first to date to directly compare sexually maltreated individuals to non-sexually maltreated individuals with matched trauma loads. This comparison enabled us to detect unique CSA effects, as differences are likely to arise from differences in *qualitative* maltreatment experiences, while controlling for trauma load, age, sex, and diagnosis. We further investigated whether both groups showed distinct alterations in brain morphology when compared to non-maltreated individuals. Our findings add new, important insights to the extant literature by providing a large sample with matched groups.

We identified a unique neural signature of childhood sexual abuse, evident in larger cerebellar GMV and widespread greater cortical thickness in frontoparietal brain areas compared to non-sexually maltreated individuals in a population with and without depression. Importantly, while cortical thickness findings were widespread, the effect sizes for both cortical thickness and GMV were small.

Regarding gray matter volume, larger cerebellar GMV distinguished *CSA* from *nCSA*, but no significant group differences were found when comparing the maltreated groups to the non-maltreated groups. This observation suggests that while there are discernible differences in cerebellar GMV when comparing *CSA* to *nCSA*, these changes do not extend to a broader contrast with a non-maltreated control group. Therefore, any conclusions about the specific impacts of non-sexual maltreatment on cerebellar volume must be tempered by the understanding that these effects might not be as pronounced or detectable when compared to a general control population with comparable age, sex distribution and diagnosis status.

The cerebellum has been implicated in social, cognitive, and emotional processes in addition to motor control (47–50). It modulates movement by regulating force, rate, rhythm, and accuracy via error-based learning (47,51–55). Given the homogeneous cytoarchitecture of the cerebellum, this error-predictive function might also extend to cognitive and emotional domains (47). Structural and functional alterations in the cerebellum have been associated with a general vulnerability for multiple mental disorders and general psychopathology (56–62). Previous studies have identified functional cerebro-cerebellar connections, especially those involved in fear and anxiety networks, including the anterior cingulate cortex (ACC), insula, inferior frontal gyrus, amygdala, hippocampus, raphe nuclei, and ventral tegmental area (63–70). Consequently, trauma-induced cerebellar alterations may set the ground for a variety of psychiatric disorders.

Several studies have described morphometric cerebellar alterations in association with CM with distinct findings in specific subgroups (e.g., psychiatric diagnosis, CM subtype, children vs. adults) (71–73). Interestingly, two studies indicate that trauma involving extreme physical threat is associated with *larger* cerebellar volume: female rape survivors with PTSD exhibited increased gray matter density in the left cerebellum, and larger cerebellar GMV was detected in combat-exposed male soldiers (74,75). Our findings of larger cerebellar volume align with these results and point to a possible neural signature of larger cerebellar volume associated with physical threat trauma types.

Compared to the *nCM* group, *CSA* showed uniquely larger GMV in the bilateral anterior cingulate gyrus and bilateral superior medial frontal gyrus. Similarly, the *nCSA* > *nCM* comparison revealed larger GMV in similar regions, which strengthens our assumption of a general trauma effect, rather than a CSA-specific effect. These findings do not align with the current meta-analysis on structural CM effects, which reported no converging evidence for regions with larger GMV in CM survivors (1). Results from our primarily female, high CM load, and largely depressed sample may reveal unique alterations that do not generalize across all survivors of CM. Of note, in a study of maltreated individuals with and without MDD, *larger* volume in the maltreated group in the right dorsomedial prefrontal cortex was reported, aligning with our results (76).

Examining cortical thickness, larger cortical thickness compared to *nCSA* was found in areas encompassing the pre- and postcentral gyrus, the anterior cingulate gyrus, insula, precuneus, superiorfrontal, superiorparietal, and superiortemporal gyri. The findings can further be contextualized using the *CSA* > *nCM* comparison. *CSA* showed similar findings when compared to *nCM,* with overlap in all these clusters. These results highlight the unique neural correlates of CSA compared to *nCSA* and *nCM*, apparent in larger cortical thickness in widespread frontoparietal regions.

*Smaller* cortical thickness is commonly associated with increased psychopathology. In MDD specifically, smaller cortical thickness was reported in the bilateral medial orbitofrontal cortex, fusiform gyrus, insula, rostral anterior and posterior cingulate cortex and in the left middle temporal gyrus, right inferior temporal gyrus, and right caudal ACC (77). As our sample, including the *nCM* group, mainly consisted of depressed individuals, cortical thinning should be the present in all three groups (*CSA, nCSA, nCM*). Consequently, our findings possibly suggest a *preserved* cortical thickness in *CSA* compared to *nCSA* and *nCM*. However, the preserved cortical thickness in CSA does not translate to reduced psychopathology, as indicated by comparable MDD duration and illness onset in the *CSA* and *nCSA* groups.

When adjusting for depression severity using HAMD, differences in cortical thickness between the *CSA* and *nCSA* groups became less pronounced in some regions (right insula, right precentral gyrus and left precuneus and cuneus), whereas the differences between *CSA* and *nCM* were more clearly defined (See Supplementary Figure 3 and Supplementary Table 10 and 11). This suggests that while depression severity modulates some of the neural impacts of CSA, particularly when compared with other forms of maltreatment, the core neurobiological alterations associated with *CSA* remain distinct and robust against a control group without maltreatment. These results underscore the importance of considering both the specific type of maltreatment and the role of concurrent psychiatric symptoms in understanding their differential impacts on brain structure.

Our findings do not align with previous findings reporting smaller cortical thickness in the somatosensory cortex in a sample of younger women with CSA (26). This difference is likely due to variations in sample characteristics, including age, as well as sample sizes and different analytical approach (11). Importantly, while previous studies have primarily focused on younger adults where trauma occurred more recently, our study examines a middle-aged sample with a mean age of 41.5 years. This represents a significant and necessary expansion of previous research, demonstrating that even decades after the trauma, CSA continues to exert measurable effects on the brain. Our findings highlight the value of studying later-life populations, as they provide unique insights into the prolonged impacts of CSA on brain over time (11).

To summarize, this study advances our understanding of the unique morphometric effects of CSA. First, our sample represents the full range of affected individuals, including healthy and depressed men and women with an extensive age range. Second, we have provided the largest study sample so far, including 195 individuals with a history of CSA. Third, we have implemented a methodologically novel approach, by matching groups according to trauma load, age, sex, and MDD diagnosis distribution.

Some limitations regarding our study design must be noted. While the rigorous matching procedure allowed us to investigate the specific effects of CSA exposure, it naturally led to an artificially high number of women and MDD patients in the *nCSA* and *nCM* comparison groups. Moreover, it resulted in a relatively high trauma load in the *nCSA* group. Together, these matched groups generally impede direct comparability to previous findings. Owing to our sample characteristics, we focused on individuals with and without MDD. It is important to note that *CSA* is also associated with higher rates of other psychiatric illnesses, such as anxiety, PTSD, and substance misuse (6). Future studies should investigate effects of CM in these populations to provide a more comprehensive understanding of the impacts of CSA and differential psychopathology on brain morphometry.

Retrospective self-report measures for CM may be influenced by recall bias, with studies indicating differences between prospective and retrospective reports (78,79). Nevertheless, the CTQ has shown good temporal stability (80). Recent meta-analytic findings suggest that retrospective measures exhibit a stronger association with psychopathology than prospective measures (81). Further, the CTQ does not assess the age at which maltreatment occurred, preventing the analysis of timing-specific effects that have been linked to varying brain outcomes in several studies (10,16,82).

Moreover, as with all cross-sectional designs, we cannot infer causality. It is possible that, as discussed by Blithikori et al. (2022) trauma-related morphometric changes are secondary to biological, psychological, or environmental factors that are not caused by, but rather associated with, CSA (57).

Our findings, using a carefully matched group design, indicate a difference in sexual vs. non-sexual abuse apparent in cortical thickness in widespread fronto-parietal areas and larger cerebellar GMV, even when controlling for general trauma load. While these brain changes may relate to adverse outcomes in the long-term, they may *initially* represent an adaptive response to highly adverse environments and a short-term survival strategy which are specific to the type of trauma experienced (4). Future studies should use longitudinal designs to further explore the impact of CSA on structural and functional connectivity, and their association with behavioral phenotypes. Given the pronounced effects of CM, and CSA specifically, mental health initiatives should focus on large-scale preventative measures and foster protective factors that can mitigate the deleterious effects of CM.

## Data Availability

All data produced in the present study are available upon reasonable request to the authors

## CRediT authorship contribution statement

**KB, VH:** Conceptualization, Data curation, Formal Analysis, Investigation, Methodology, Project Administration, Writing – original draft. **PU, FSt**, **SZ**, **FSch**, **FTO**, **LT**, **SM**, **KT**, **KF**, **NS**, **JG**, **EL, LB**, **DG, NW, ED:** Data curation, Writing – review & editing. **TH**, **BS**, **HJ**, **AJ**, **AK**, **DU**, **IN, TK:** Funding Acquisition, Writing – review & editing. **NA:** Conceptualization, Funding Acquisition, Methodology, Supervision, Writing – review & editing.

## Acknowledgements

The FOR2107 cohort project was approved by the Ethics Committees of the Medical Faculties, University of Marburg (AZ:07/14) and University of Münster (AZ:2014-422-b-S).

We are deeply indebted to all study participants and staff. A list of acknowledgments can be found here: https://for2107.de/acknowledgements/?lang=en.

## References

1. Yang W, Jin S, Duan W, Yu H, Ping L, Shen Z, et al. The effects of childhood maltreatment on cortical thickness and gray matter volume: a coordinate-based meta-analysis. Psychol Med [Internet]. 2023 Mar 22 [cited 2023 Sep 22];53(5). Available from: https://pubmed.ncbi.nlm.nih.gov/36946124/

2. Madigan S, Deneault AA, Racine N, Park J, Thiemann R, Zhu J, et al. Adverse childhood experiences: a meta-analysis of prevalence and moderators among half a million adults in 206 studies. World Psychiatry [Internet]. 2023 Oct 1 [cited 2024 Jan 10];22(3):463–71. Available from: https://onlinelibrary.wiley.com/doi/full/10.1002/wps.21122

3. Gallo EAG, Munhoz TN, Loret de Mola C, Murray J. Gender differences in the effects of childhood maltreatment on adult depression and anxiety: A systematic review and meta-analysis. Child Abuse Negl. 2018;79(March 2017):107–14.

4. Teicher MH, Samson JA, Anderson CM, Ohashi K. The effects of childhood maltreatment on brain structure, function and connectivity. Nat Rev Neurosci [Internet]. 2016;17(10):652–66. Available from: 10.1038/nrn.2016.111

5. McLaughlin KA, Sheridan MA, Lambert HK. Childhood Adversity and Neural Development: Deprivation and Threat as Distinct Dimensions of Early Experience NIH Public Access. Neurosci Biobehav Rev. 2014;47:578–91.

6. Hailes HP, Yu R, Danese A, Fazel S. Long-term outcomes of childhood sexual abuse: an umbrella review. Lancet Psychiatry. 2019 Oct 1;6(10):830–9.

7. World Health Organisation. https://www.who.int/news-room/fact-sheets/detail/child-maltreatment. 2022. Child Maltreatment.

8. Putnam FW. Ten-Year Research Update Review: Child Sexual Abuse. J Am Acad Child Adolesc Psychiatry. 2003 Mar 1;42(3):269–78.

9. Afifi TO, MacMillan HL, Boyle M, Taillieu T, Cheung K, Sareen J. Child abuse and mental disorders in Canada. CMAJ [Internet]. 2014 Jun 10 [cited 2023 Jul 25];186(9). Available from: https://pubmed.ncbi.nlm.nih.gov/24756625/

10. Spataro J, Mullen PE, Burgess PM, Wells DL, Moss SA. Impact of child sexual abuse on mental health: prospective study in males and females. Br J Psychiatry [Internet]. 2004 May [cited 2023 Aug 30];184(MAY):416–21. Available from: https://pubmed.ncbi.nlm.nih.gov/15123505/

11. Paquola C, Bennett MR, Lagopoulos J. Understanding heterogeneity in grey matter research of adults with childhood maltreatment—A meta-analysis and review. Neurosci Biobehav Rev [Internet]. 2016;69:299–312. Available from: 10.1016/j.neubiorev.2016.08.011

12. Brown M, Worrell C, Pariante CM. Inflammation and early life stress: An updated review of childhood trauma and inflammatory markers in adulthood. Pharmacol Biochem Behav. 2021 Dec 1;211:173291.

13. Hosseini-Kamkar N, Varvani Farahani M, Nikolic M, Stewart K, Goldsmith S, Soltaninejad M, et al. Adverse Life Experiences and Brain Function: A Meta-Analysis of Functional Magnetic Resonance Imaging Findings. JAMA Netw Open [Internet]. 2023 Nov 1 [cited 2024 Feb 11];6(11):e2340018–e2340018. Available from: https://jamanetwork.com/journals/jamanetworkopen/fullarticle/2811174

14. Myroniuk S, Reitsema AM, De Jonge P, Jeronimus BF. Childhood abuse and neglect and profiles of adult emotion dynamics. Dev Psychopathol [Internet]. 2024 [cited 2024 Feb 11];1–19. Available from: https://www.cambridge.org/core/journals/development-and-psychopathology/article/childhood-abuse-and-neglect-and-profiles-of-adult-emotion-dynamics/0F7161ABCDED2D2887E37388E5E539BA

15. McTavish JR, Santesso N, Amin A, Reijnders M, Ali MU, Fitzpatrick-Lewis D, et al. Psychosocial interventions for responding to child sexual abuse: A systematic review. Child Abuse Negl. 2021 Jun 1;116:104203.

16. Li M, Gao T, Su Y, Zhang Y, Yang G, D’Arcy C, et al. The Timing Effect of Childhood Maltreatment in Depression: A Systematic Review and meta-Analysis. 10.1177/15248380221102558 [Internet]. 2022 May 24 [cited 2024 May 12];24(4):2560–80. Available from: https://journals.sagepub.com/doi/10.1177/15248380221102558?url_ver=Z39.88-2003&rfr_id=ori%3Arid%3Acrossref.org&rfr_dat=cr_pub++0pubmed

17. Goltermann J, Winter N, Meinert S, Grotegerd D, Kraus A, Flinkenflügel K, et al. Gray matter correlates of childhood maltreatment in the context of major depression: searching for replicability in a multi-cohort brain-wide association study of 3225 adults. bioRxiv [Internet]. 2024 Aug 16 [cited 2024 Dec 29];2024.08.15.608132. Available from: https://www.biorxiv.org/content/10.1101/2024.08.15.608132v1

18. Blanco L, Nydegger LA, Camarillo G, Trinidad DR, Schramm E, Ames SL. Neurological changes in brain structure and functions among individuals with a history of childhood sexual abuse: A review. Neurosci Biobehav Rev [Internet]. 2015 Oct 1 [cited 2024 May 12];57:63–9. Available from: https://pubmed.ncbi.nlm.nih.gov/26363666/

19. Andersen SL, Tomada A, Vincow ES, Valente E, Polcari A, Teicher MH. Preliminary evidence for sensitive periods in the effect of childhood sexual abuse on regional brain development. J Neuropsychiatry Clin Neurosci [Internet]. 2008 [cited 2024 May 12];20(3):292–301. Available from: https://pubmed.ncbi.nlm.nih.gov/18806232/

20. Stein MB, Koverola C, Hanna C, Torchia MG, McClarty B. Hippocampal volume in women victimized by childhood sexual abuse. Psychol Med [Internet]. 1997 Jul [cited 2024 May 12];27(4):951–9. Available from: https://pubmed.ncbi.nlm.nih.gov/9234472/

21. Teicher MH, Dumont NL, Ito Y, Vaituzis C, Giedd JN, Andersen SL. Childhood neglect is associated with reduced corpus callosum area. Biol Psychiatry [Internet]. 2004 Jul 15 [cited 2024 May 12];56(2):80–5. Available from: https://pubmed.ncbi.nlm.nih.gov/15231439/

22. Vythilingam M, Heim C, Newport J, Miller AH, Anderson E, Bronen R, et al. Childhood Trauma Associated With Smaller Hippocampal Volume in Women With Major Depression. Am J Psychiatry [Internet]. 2002 Dec 1 [cited 2024 May 12];159(12):2072. Available from: /pmc/articles/PMC3230324/

23. Bremner JD, Vythilingam M, Vermetten E, Southwick SM, McGlashan T, Nazeer A, et al. MRI and PET study of deficits in hippocampal structure and function in women with childhood sexual abuse and posttraumatic stress disorder. Am J Psychiatry [Internet]. 2003 May [cited 2024 May 12];160(5):924–32. Available from: https://pubmed.ncbi.nlm.nih.gov/12727697/

24. Rinne-Albers MA, Boateng CP, van der Werff SJ, Lamers-Winkelman F, Rombouts SA, Vermeiren RR, et al. Preserved cortical thickness, surface area and volume in adolescents with PTSD after childhood sexual abuse. Sci Rep [Internet]. 2020 Dec 1 [cited 2024 Feb 4];10(1). Available from: https://pubmed.ncbi.nlm.nih.gov/32094427/

25. Kim J, Lee C, Kang Y, Kang W, Kim A, Tae WS, et al. Childhood Sexual Abuse and Cortical Thinning in Adults With Major Depressive Disorder. Psychiatry Investig [Internet]. 2023 Mar 1 [cited 2024 Jan 11];20(3):255–61. Available from: http://psychiatryinvestigation.org/journal/view.php?doi=10.30773/pi.2022.0314

26. Heim CM, Mayberg HS, Mletzko T, Nemeroff CB, Pruessner JC. Decreased cortical representation of genital somatosensory field after childhood sexual abuse. American Journal of Psychiatry. 2013;170(6):616–23.

27. Tomoda A, Navalta CP, Polcari A, Sadato N, Teicher MH. Childhood Sexual Abuse Is Associated with Reduced Gray Matter Volume in Visual Cortex of Young Women. Biol Psychiatry [Internet]. 2009;66(7):642–8. Available from: 10.1016/j.biopsych.2009.04.021

28. Kim SY, An SJ, Han JH, Kang Y, Bae EB, Tae WS, et al. Childhood abuse and cortical gray matter volume in patients with major depressive disorder. Psychiatry Res. 2023 Jan 1;319:114990.

29. Yu M, Cullen N, Linn KA, Oathes DJ, Seok D, Cook PA, et al. Structural brain measures linked to clinical phenotypes in major depression replicate across clinical centres. Molecular Psychiatry 2021 26:7 [Internet]. 2021 Feb 15 [cited 2024 Feb 4];26(7):2764–75. Available from: https://www.nature.com/articles/s41380-021-01039-8

30. Walker EA, Gelfand A, Katon WJ, Koss MP, Von Korff M, Bernstein D, et al. Adult health status of women with histories of childhood abuse and neglect. American Journal of Medicine. 1999;107(4):332–9.

31. Felitti VJ, Anda RF, Nordenberg D, Williamson DF, Spitz AM, Edwards V, et al. Relationship of childhood abuse and household dysfunction to many of the leading causes of death in adults: The adverse childhood experiences (ACE) study. Am J Prev Med [Internet]. 1998 May [cited 2023 Jul 25];14(4):245–58. Available from: https://pubmed.ncbi.nlm.nih.gov/9635069/

32. Teicher MH, Samson JA. Childhood maltreatment and psychopathology: A case for ecophenotypic variants as clinically and neurobiologically distinct subtypes. American Journal of Psychiatry. 2013;170(10):1114–33.

33. Tozzi L, Garczarek L, Janowitz D, Stein DJ, Wittfeld K, Dobrowolny H, et al. Interactive impact of childhood maltreatment, depression, and age on cortical brain structure: Mega-analytic findings from a large multi-site cohort. Psychol Med. 2020;50(6):1020–31.

34. Brosch K, Stein F, Meller T, Schmitt S, Yuksel D, Ringwald KG, et al. DLPFC volume is a neural correlate of resilience in healthy high-risk individuals with both childhood maltreatment and familial risk for depression. Psychol Med. 2021 Apr;1–7.

35. Kircher T, Wöhr M, Nenadic I, Schwarting R, Schratt G, Alferink J, et al. Neurobiology of the major psychoses: a translational perspective on brain structure and function-the FOR2107 consortium. Eur Arch Psychiatry Clin Neurosci. 2018;1:3.

36. Wittchen HU, Gruschwitz S, Wunderlich U, Zaudig M. Strukturiertes Klinisches Interview für DSM-IV (SKID-I). Achse I: Psychische Störungen. Göttingen: Hogrefe. 1997.

37. Lehrl S, Triebig G, Fischer B. Multiple choice vocabulary test MWT as a valid and short test to estimate premorbid intelligence. Acta Neurol Scand. 1995 May;91(5):335–45.

38. Wingenfeld K, Spitzer C, Mensebach C, Grabe HJ, Hill A, Gast U, et al. The german version of the Childhood Trauma Questionnaire (CTQ):Preliminary psychometric properties. PPmP Psychotherapie Psychosomatik Medizinische Psychologie. 2010;60(11):442–50.

39. Vogelbacher C, Möbius TWD, Sommer J, Schuster V, Dannlowski U, Kircher T, et al. The Marburg-Münster Affective Disorders Cohort Study (MACS): A quality assurance protocol for MR neuroimaging data. Neuroimage. 2018;172(January):450–60.

40. Ashburner J. A fast diffeomorphic image registration algorithm. Neuroimage. 2007 Oct 15;38(1):95–113.

41. Ho DE, Imai K, King G, Stuart EA. Matching as nonparametric preprocessing for reducing model dependence in parametric causal inference. Political Analysis. 2007;15(3):199–236.

42. Smith SM, Nichols TE. Threshold-free cluster enhancement: addressing problems of smoothing, threshold dependence and localisation in cluster inference. Neuroimage [Internet]. 2009 Jan 1 [cited 2023 Oct 16];44(1):83–98. Available from: https://pubmed.ncbi.nlm.nih.gov/18501637/

43. Desikan RS, Ségonne F, Fischl B, Quinn BT, Dickerson BC, Blacker D, et al. An automated labeling system for subdividing the human cerebral cortex on MRI scans into gyral based regions of interest. Neuroimage. 2006 Jul 1;31(3):968–80.

44. Teicher MH, Gordon JB, Nemeroff CB. Recognizing the importance of childhood maltreatment as a critical factor in psychiatric diagnoses, treatment, research, prevention, and education. Molecular Psychiatry 2021 27:3 [Internet]. 2021 Nov 4 [cited 2024 Feb 7];27(3):1331–8. Available from: https://www.nature.com/articles/s41380-021-01367-9

45. Warrier V, Kwong ASF, Luo M, Dalvie S, Croft J, Sallis HM, et al. Gene–environment correlations and causal effects of childhood maltreatment on physical and mental health: a genetically informed approach. Lancet Psychiatry [Internet]. 2021 May 1 [cited 2024 Feb 7];8(5):373–86. Available from: http://www.thelancet.com/article/S2215036620305691/fulltext

46. Schmaal L, Hibar DP, Sämann PG, Hall GB, Baune BT, Jahanshad N, et al. Cortical abnormalities in adults and adolescents with major depression based on brain scans from 20 cohorts worldwide in the ENIGMA Major Depressive Disorder Working Group. Mol Psychiatry. 2017;22(6):900–9.

47. Schmahmann JD, Guell X, Stoodley CJ, Halko MA. The Theory and Neuroscience of Cerebellar Cognition. 2019 [cited 2023 Oct 16]; Available from: 10.1146/annurev-neuro-070918-

48. Van Overwalle F, Manto M, Cattaneo Z, Clausi S, Ferrari C, Gabrieli JDE, et al. Consensus Paper: Cerebellum and Social Cognition. Cerebellum [Internet]. 2020 Dec 1 [cited 2023 Oct 15];19(6):833–68. Available from: https://link.springer.com/article/10.1007/s12311-020-01155-1

49. Krautheim JT, Steines M, Dannlowski U, Neziroğlu G, Acosta H, Sommer J, et al. Emotion specific neural activation for the production and perception of facial expressions. Cortex [Internet]. 2020 Jun 1 [cited 2024 Aug 4];127:17–28. Available from: https://pubmed.ncbi.nlm.nih.gov/32155474/

50. Steines M, Krautheim JT, Neziroğlu G, Kircher T, Straube B. Conflicting group memberships modulate neural activation in an emotional production-perception network. Cortex [Internet]. 2020 May 1 [cited 2024 Aug 4];126:153–72. Available from: https://pubmed.ncbi.nlm.nih.gov/32078820/

51. Ito M. Historical Review of the Significance of the Cerebellum and the Role of Purkinje Cells in Motor Learning. Ann N Y Acad Sci [Internet]. 2002 Dec 1 [cited 2023 Oct 16];978(1):273–88. Available from: https://onlinelibrary.wiley.com/doi/full/10.1111/j.1749-6632.2002.tb07574.x

52. ten Brinke MM, Heiney SA, Wang X, Proietti-Onori M, Boele HJ, Bakermans J, et al. Dynamic modulation of activity in cerebellar nuclei neurons during pavlovian eyeblink conditioning in mice. [cited 2023 Oct 16]; Available from: 10.7554/eLife.28132.001

53. Heffley W, Hull C. Classical conditioning drives learned reward prediction signals in climbing fibers across the lateral cerebellum. 2019 [cited 2023 Oct 16]; Available from: 10.7554/eLife.46764.001

54. Hull C. Prediction signals in the cerebellum: Beyond supervised motor learning. Elife. 2020 Mar 1;9.

55. Sokolov AA, Miall RC, Ivry RB. The Cerebellum: Adaptive Prediction for Movement and Cognition. Trends Cogn Sci. 2017 May 1;21(5):313–32.

56. Talati A, Pantazatos SP, Schneier FR, Weissman MM, Hirsch J. Grey Matter Abnormalities in Social Anxiety Disorder: Primary, Replication, and Specificity Studies. 2012;

57. Blithikioti C, Nuño L, Guell X, Pascual-Diaz S, Gual A, Balcells-Olivero, et al. The cerebellum and psychological trauma: A systematic review of neuroimaging studies. Neurobiol Stress [Internet]. 2022 Mar 1 [cited 2023 Sep 19];17:100429. Available from: /pmc/articles/PMC8801754/

58. Romer AL, Knodt AR, Houts R, Brigidi BD, Moffitt TE, Caspi A, et al. Structural alterations within cerebellar circuitry are associated with general liability for common mental disorders. Mol Psychiatry. 2018;23(4):1084–90.

59. Amaral DG, Schumann CM, Nordahl CW. Neuroanatomy of autism. Trends Neurosci. 2008 Mar 1;31(3):137–45.

60. Shinn AK, Roh YS, Ravichandran CT, Baker JT, Öngür D, Cohen BM. Aberrant cerebellar connectivity in bipolar disorder with psychosis. Biol Psychiatry Cogn Neurosci Neuroimaging [Internet]. 2017 Jul 1 [cited 2023 Oct 16];2(5):438–48. Available from: https://pubmed.ncbi.nlm.nih.gov/28730183/

61. Moberget T, Doan NT, Alnæs D, Kaufmann T, Córdova-Palomera A, Lagerberg T V., et al. Cerebellar volume and cerebellocerebral structural covariance in schizophrenia: a multisite mega-analysis of 983 patients and 1349 healthy controls. Molecular Psychiatry 2018 23:6 [Internet]. 2017 May 16 [cited 2023 Oct 16];23(6):1512–20. Available from: https://www.nature.com/articles/mp2017106

62. Hariri AR. The Emerging Importance of the Cerebellum in Broad Risk for Psychopathology. Neuron [Internet]. 2019 Apr 3 [cited 2024 Jun 13];102(1):17–20. Available from: https://pubmed.ncbi.nlm.nih.gov/30946819/

63. Addis DR, Moloney EEJ, Tippett LJ, P. Roberts R, Hach S. Characterizing cerebellar activity during autobiographical memory retrieval: ALE and functional connectivity investigations. Neuropsychologia. 2016 Sep 1;90:80–93.

64. Etkin A, Wager TD. Functional Neuroimaging of Anxiety: A Meta-Analysis of Emotional Processing in PTSD, Social Anxiety Disorder, and Specific Phobia. Am J Psychiatry [Internet]. 2007 Oct [cited 2023 Oct 16];164(10):1476. Available from: /pmc/articles/PMC3318959/

65. Beliveau V, Svarer C, Frokjaer VG, Knudsen GM, Greve DN, Fisher PM. Functional connectivity of the dorsal and median raphe nuclei at rest. Neuroimage. 2015 Aug 1;116:187–95.

66. Cservenka A, Casimo K, Fair DA, Nagel BJ. Resting state functional connectivity of the nucleus accumbens in youth with a family history of alcoholism. Psychiatry Res Neuroimaging. 2014 Mar 30;221(3):210–9.

67. Hwang KD, Kim SJ, Lee YS. Cerebellar Circuits for Classical Fear Conditioning. Front Cell Neurosci. 2022 Mar 30;16:836948.

68. Moreno-Rius J. The cerebellum in fear and anxiety-related disorders. Prog Neuropsychopharmacol Biol Psychiatry. 2018 Jul 13;85:23–32.

69. Sang L, Qin W, Liu Y, Han W, Zhang Y, Jiang T, et al. Resting-state functional connectivity of the vermal and hemispheric subregions of the cerebellum with both the cerebral cortical networks and subcortical structures. Neuroimage. 2012 Jul 16;61(4):1213–25.

70. Zeng LL, Shen H, Liu L, Wang L, Li B, Fang P, et al. Identifying major depression using whole-brain functional connectivity: a multivariate pattern analysis. Brain [Internet]. 2012 May 1 [cited 2023 Oct 16];135(5):1498–507. Available from: 10.1093/brain/aws059

71. Pollok TM, Kaiser A, Kraaijenvanger EJ, Monninger M, Brandeis D, Banaschewski T, et al. Neurostructural traces of early life adversities: A meta-analysis exploring age- and adversity-specific effects. Neurosci Biobehav Rev [Internet]. 2022 Apr 1 [cited 2023 Oct 16];135:104589. Available from: /pmc/articles/PMC9013474/

72. Gehred MZ, Knodt AR, Ambler A, Bourassa KJ, Danese A, Elliott ML, et al. Long-term Neural Embedding of Childhood Adversity in a Population-Representative Birth Cohort Followed for 5 Decades. Biol Psychiatry [Internet]. 2021 Aug 8 [cited 2023 Oct 16];90(3):182. Available from: /pmc/articles/PMC8274314/

73. Walsh ND, Dalgleish T, Lombardo M V., Dunn VJ, Van Harmelen AL, Ban M, et al. General and specific effects of early-life psychosocial adversities on adolescent grey matter volume. Neuroimage Clin. 2014 Jan 1;4:308–18.

74. Sui SG, Zhang Y, Wu MX, Xu JM, Duan L, Weng XC, et al. Abnormal cerebellum density in victims of rape with post-traumatic stress disorder: Voxel-based analysis of magnetic resonance imaging investigation. Asia-Pacific Psychiatry. 2010;2(3).

75. Sussman D, Pang EW, Jetly R, Dunkley BT, Taylor MJ. Neuroanatomical features in soldiers with post-traumatic stress disorder. BMC Neurosci [Internet]. 2016 [cited 2023 Oct 16];17(1):13. Available from: /pmc/articles/PMC4815085/

76. Chaney A, Carballedo A, Amico F, Fagan A, Skokauskas N, Meaney J, et al. Effect of childhood maltreatment on brain structure in adult patients with major depressive disorder and healthy participants. J Psychiatry Neurosci [Internet]. 2014 [cited 2023 Oct 15];39(1):50. Available from: /pmc/articles/PMC3868665/

77. Schmaal L, Veltman DJ, Van Erp TGM, Smann PG, Frodl T, Jahanshad N, et al. Subcortical brain alterations in major depressive disorder: Findings from the ENIGMA Major Depressive Disorder working group. Mol Psychiatry. 2016;21(6):806–12.

78. Baldwin JR, Reuben A, Newbury JB, Danese A. Agreement Between Prospective and Retrospective Measures of Childhood Maltreatment: A Systematic Review and Meta-analysis. JAMA Psychiatry [Internet]. 2019 Jun 1 [cited 2024 Feb 11];76(6):584–93. Available from: https://jamanetwork.com/journals/jamapsychiatry/fullarticle/2728182

79. Newbury JB, Arseneault L, Moffitt TE, Caspi A, Danese A, Baldwin JR, et al. Measuring childhood maltreatment to predict early-adult psychopathology: Comparison of prospective informant-reports and retrospective self-reports. J Psychiatr Res. 2018 Jan 1;96:57–64.

80. Goltermann J, Meinert S, Hülsmann C, Dohm K, Grotegerd D, Redlich R, et al. Temporal stability and state-dependence of retrospective self-reports of childhood maltreatment in healthy and depressed adults. Psychol Assess [Internet]. 2023 Nov 10 [cited 2024 Feb 11];35(1):12–22. Available from: https://pubmed.ncbi.nlm.nih.gov/36355690/

81. Baldwin JR, Coleman O, Francis ER, Danese A. Prospective and Retrospective Measures of Child Maltreatment and Their Association With Psychopathology: A Systematic Review and Meta-Analysis. JAMA Psychiatry [Internet]. 2024 [cited 2024 Jun 11]; Available from: https://pubmed.ncbi.nlm.nih.gov/38691376/

82. Holz NE, Zabihi M, Kia SM, Monninger M, Aggensteiner PM, Siehl S, et al. A stable and replicable neural signature of lifespan adversity in the adult brain. Nature Neuroscience 2023 26:9 [Internet]. 2023 Aug 21 [cited 2024 Jun 13];26(9):1603–12. Available from: https://www.nature.com/articles/s41593-023-01410-8

