## Supplement for "Unique neural signatures of childhood sexual abuse revisited"

**Supplementary Figure 1:** Cerebellar volume eigenvalues extracted from the *CSA* > *nCSA* comparison by group

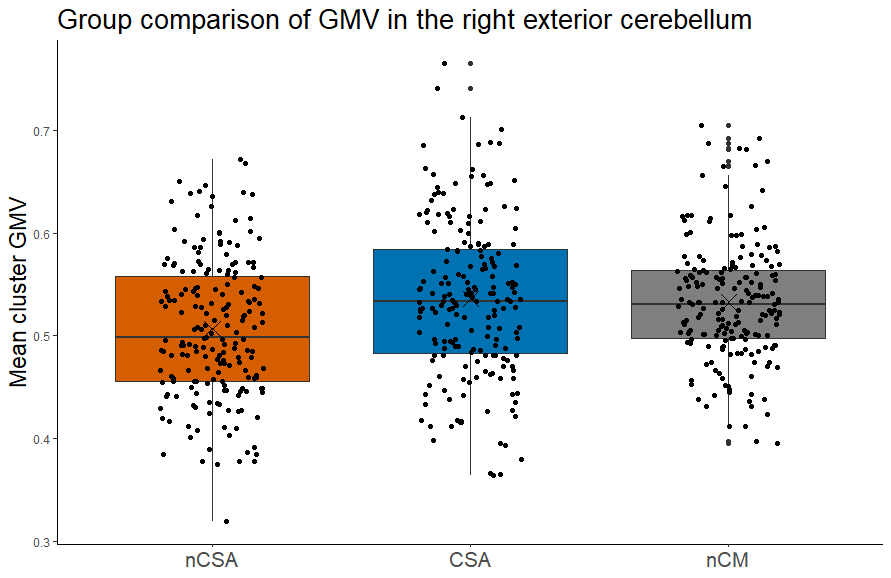

*Note*: Groups: *CSA* = Childhood sexual abuse, *nCSA* = other types of childhood maltreatment, but no sexual abuse, *nCM* = no experiences of childhood maltreatment. *CSA* and *nCSA* differ significantly in their GMV in the cerebellar cluster (*k*=1225, TFCE=917.92, *p*_FWE_=0.025, x/y/z=33/-48/-32).

**Supplementary Figure 2:** Clusters exhibiting significantly larger GMV (TFCE, *p*_FWE_-corrected) in **A** *CSA* > *nCSA* (red) and **B** *CSA* > *nCM* (red) and *nCSA* > *nCM* (blue), **controlling for comorbidity (yes/no)**.

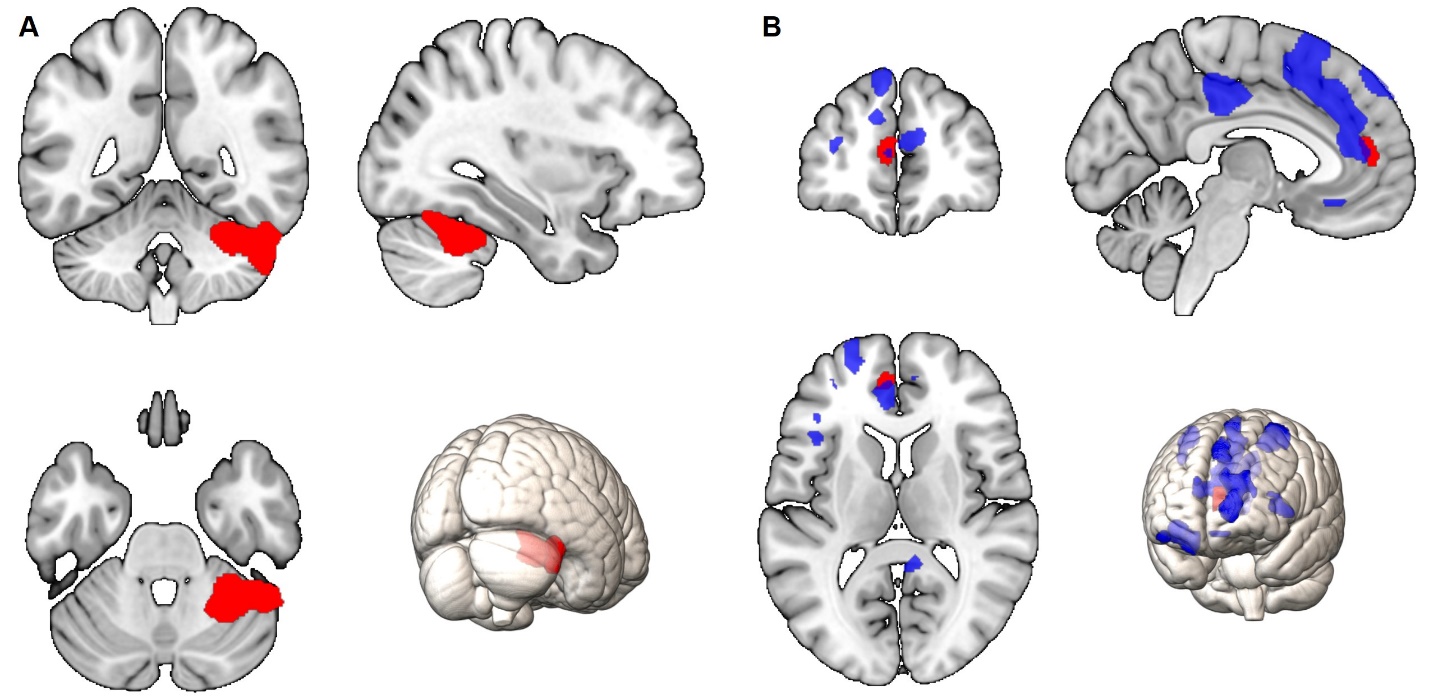

*Note.* **A** *CSA* > *nCSA* group comparison, significant cluster encompasses parts of the right exterior cerebellum (*k*=2443, TFCE=930.61, *p*­_FWE_=0.012, x/y/z=33/-48/-30), red cluster. **B** *CSA* > *nCM* group comparison; significant cluster comprises parts of the left anterior cingulate gyrus and left superior medial frontal gyrus (*k*=217, TFCE=1170.25, *p*­_FWE_=0.038, x/y/z=-04/46/10), red cluster. *nCSA* > *nCM* group comparison; significant clusters include the bilateral superior frontal gyri, middle frontal gyri, precentral gyri, orbital gyri and cingulate gyri (Cluster 1: *k*=7741, TFCE=1516.54, *p*_FWE_=0.010, x/y/z=-06/21/62; Cluster 2: *k*=1244, TFCE=1344.70, *p*_FWE_= 0.018, x/y/z=-06/-27/38; Cluster 3: *k*=1135, TFCE=1278.78, *p*_FWE_=0.023, x/y/z=-28/0/38; Cluster 4: *k*=443, TFCE=1200.67, *p*_FWE_=0.031, x/y/z=33/-04/60; Cluster 5: *k*=731, TFCE=1183.45, *p*_FWE_=0.033, x/y/z=30/38/-09; Cluster 6: *k*=543, TFCE=1109.49, *p*_FWE_=0.042, x/y/z=08/-46/18; Cluster 7: *k*=287, TFCE=1105.45, *p*_FWE_=0.043, x/y/z=-48/20/0), blue clusters. Findings show larger GMV in the respective clusters in the *CSA* group compared to *nCSA* and *nCM* (red) and in *nCSA* compared to *nCM* group (blue).

**Supplementary Figure 3:** Clusters exhibiting significantly larger cortical thickness (TFCE, *p*_FWE_-corrected) in *CSA* compared *to* ***A*** *nCSA and* ***B*** *nCM,* controlling for symptom severity (HAMD sum score)

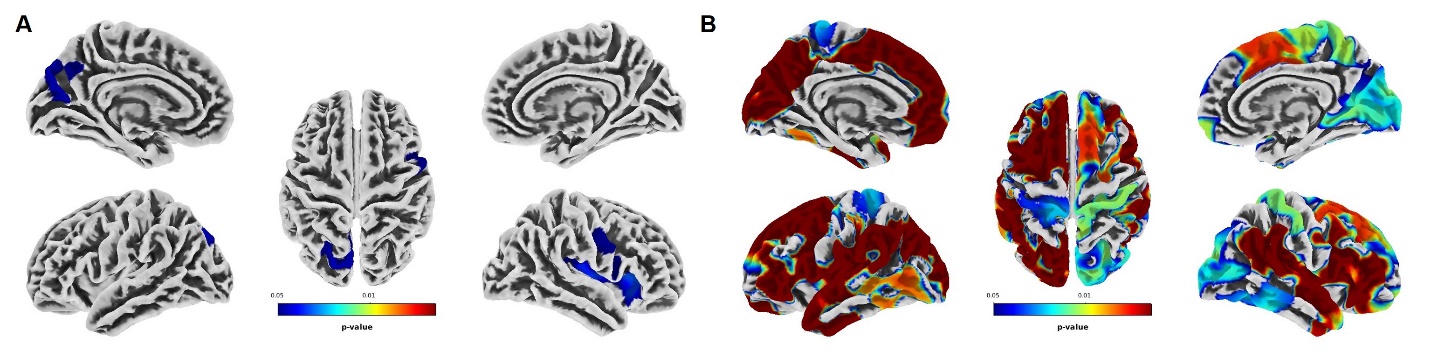

*Note.* Warmer colors represent lower *p*-values (threshold *p* < 0.05). **A** *CSA* > *nCSA* group comparison, significant clusters encompass regions of the right insula, right lateral orbitofrontal gyrus, right pars triangularis, right precentral gyrus, right pars opercularis, left superior parietal cortex, left cuneus, left precuneus and left inferior parietal gyrus. **B** *CSA* > *no CM* group comparison, significant clusters encompass regions of the bilateral superiorfrontal gyri, supramarginal gyri, superior temporal gyri, pre- and postcentral gyri, inferior- and superior parietal gyri, and the insula. See Supplementary Table 10 and 11 for a detailed list of respective significant clusters.

**Supplementary Figure 4:** Clusters exhibiting significantly larger cortical thickness (TFCE, *p*_FWE_-corrected) in *CSA* compared *to* ***A*** *nCSA and* ***B*** *nCM,* controlling for comorbidity (yes/no)

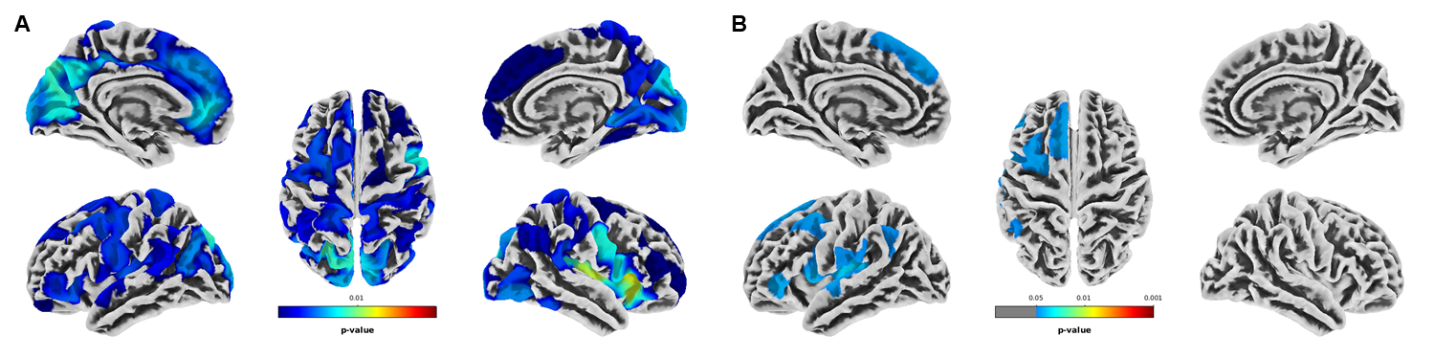

*Note.* Warmer colors represent lower *p*-values (threshold *p* < 0.05). **A** *CSA* > *nCSA* group comparison, significant clusters encompass regions of the bilateral superiorfrontal gyri, supramarginal gyri, precentral gyri, pre- and postcentral gyri, inferior- and superior parietal gyri, and the insula. **B** *CSA* > *no CM* group comparison, significant clusters encompass regions of the left caudalmiddlefrontal gyrus, left insula, left parsopercularis, left supramarginal gyrus, left inferior parietal gyrus, left pre- and postcentral gyri, and left superiortemporal gyrus. See Supplementary Table 12 and 13 for a detailed list of respective significant clusters.

***nCSA* > *nCM*; cortical thickness: no significant findings**

**Supplementary Table 1**

*Sample Characteristics (N=390)*

|  | **CSA**  (n=195) | **nCSA**  (n=195) | **Bayes Factor (BF)** |
| --- | --- | --- | --- |
| Age | 41.5 *(13.6)* | 41.8 *(13.6)* | **0.12** |
| Sex (f / m) | 168 / 27 | 164 / 31 | **0.01** |
| Diagnosis (HC / MDD) | 45 / 150 | 38 / 157 | **0.01** |
| Years of education | 12.9 *(2.6)* | 12.9 *(2.8)* | **0.11** |
| AUDIT sum score | 3.5 (3.6) | 3.4 (3.2) | **0.12** |
| IQ (estimated) | 114.5 *(13.3)* | 112.4 *(13.9)* | 0.37 |
| Household income (€) | 2183.3 (1451.6) | 2285.3 (1728.2) | **0.14** |
| MDD patients |  |  |  |
| Duration of illness (months) | 62.7 *(80.2)* | 58.6 *(90.2)* | **0.15** |
| Age of onset | 25.8 *(13.7)* | 27.3 *(13.2)* | **0.20** |
| Psychiatric medication (yes/no) | 103 / 92 | 104 / 91 | **0.01** |
| Medication load index | 1.1 (1.4) | 1.2 (1.5) | **0.12** |
| Time since treatment (months) | 101.9 (111.5) | 92.2 (91.8) | **0.18** |
| Remission (acute/partial/full) | 72 / 40 / 38 | 79 / 27 / 51 | **3.25e-3** |
| Comorbidity (yes/no) | 87 / 108 | 83 / 112 | **0.01** |
| CTQ |  |  |  |
| sum score | 57.2 *(15.9)* | 54.8 *(12.2)* | 0.41 |
| multiple traumatization (yes/no) | 160 / 35 | 156 / 39 | **0.01** |
| sexual abuse | 12.0 *(4.1)* | 5.3 *(0.6)* | **8.86e+69** |
| emotional abuse | 13.0 *(5.3)* | 14.7 *(4.9)* | **23.13** |
| physical abuse | 8.0 *(3.7)* | 8.5 *(3.9)* | **0.25** |
| emotional neglect | 14.9 *(5.4)* | 16.9 *(4.7)* | **102.47** |
| physical neglect | 9.2 *(3.7)* | 9.4 *(2.9)* | **0.14** |
| BDI sum score | 17.4 (*12.3)* | 15.7 *(11.3)* | **0.27** |
| HAMD sum score | 8.6 *(6.9)* | 7.9 *(6.8)* | **0.16** |
| PSS sum score | 28.7 *(10.1)* | 27.5 *(10.8)* | **0.21** |
| FSozU sum score | 3.8 *(0.9)* | 3.8 *(0.9)* | **0.13** |
| RS-25 sum score | 115.6 *(28.0)* | 116.9 *(28.4)* | **0.12** |
| GAF score | 68.1 *(16.2)* | 68.2 *(16.8)* | **0.11** |
| LEQ negative events score | 15.0 *(14.1)* | 15.5 *(19.0)* | **0.12** |
| LEQ positive events score | 9.0 *(8.2)* | 8.9 *(10.4)* | **0.11** |

*Note*: Mean *(standard deviation)*, AUDIT (Alcohol Use Disorders Identification Test), IQ (Multiple choice vocabulary test, MWT-B), Medication (acute psychotropic medication intake), CTQ (childhood trauma questionnaire), BDI (Beck Depression Inventory), HAMD (Hamilton rating scale for Depression, 21 item version), PSS (Perceived Stress Scale), FSozU (Perceived Social Support Questionnaire), RS-25 (Resilience Questionnaire), GAF (Global Assessment of Functioning), LEQ (Life events questionnaire). The Bayes Factor (BF) values quantify the evidence in favor of H₁ (dependence of the variable on the group) compared to H₀ (independence). BF values >1 indicate support for dependence, thus indicate evidence in favor of a group effect, while BF values <1 favor independence. The interpretation of BF values follows common thresholds: **A) BF < 0.1**: Strong evidence for independence (null hypothesis). **B)** **0.1 ≤ BF < 0.33**: Moderate evidence for independence (null hypothesis). **C)** **0.33 ≤ BF < 1** Anecdotal evidence for independence. **D)** **1 < BF < 3.2**: Anecdotal evidence for dependence. **E)** **3.2 ≤ BF < 10**: Moderate evidence for dependence. **F)** **10 < BF ≤ 100**: Strong evidence for dependence. **G)** **BF > 100**: Decisive evidence for dependence. Bold values indicate moderate or stronger evidence for independence (< 0.33) or dependence (>3.2). Bayesian ANOVA was performed to test for associations between continuous variables and group, and Bayesian contingency table analysis was conducted for categorical variables.

**Supplementary Table 2**

GMV: FWE-corrected clusters exhibiting significantly larger GMV in *CSA* > *nCSA*.

| *k* | TFCE | *p*FWE | coordinates (x/y/z) | % | anatomical region |
| --- | --- | --- | --- | --- | --- |
| 1225 | 917.92 | 0.025 | 33/-48/-32 | 77.3 | r. exterior cerebellum |
|  |  |  |  | 20.3 | r. cerebellum white matter |
|  |  |  |  | 2.4 | r. fusiform gyrus |
|  | 911.48 | 0.025 | 34/-56/-26 | 82.0 | r. exterior cerebellum |
|  |  |  |  | 17.3 | r. fusiform gyrus |
|  |  |  |  | 0.6 | r. occipital fusiform gyrus |
|  |  |  |  | 0.2 | r. cerebellum white matter |
|  | 888.14 | 0.029 | 51/-30/-30 | 44.5 | r. inferior temporal gyrus |
|  |  |  |  | 43.7 | r. exterior cerebellum |
|  |  |  |  | 8.1 | Background |
|  |  |  |  | 3.8 | r. fusiform gyrus |

**Supplementary Table 3**

GMV: FWE-corrected clusters exhibiting significantly larger GMV in *CSA* > *nCM*.

| *k* | TFCE | *p*FWE | coordinates (x/y/z) | % | anatomical region |
| --- | --- | --- | --- | --- | --- |
| 842 | 1007.25 | 0.024 | -04/46/12 | 43.7 | l. superior medial frontal gyrus |
|  |  |  |  | 42.1 | l. anterior cingulate gyrus |
|  |  |  |  | 8.7 | r. superior medial frontal gyrus |
|  |  |  |  | 5.4 | r. anterior cingulate gyrus |
|  | 940.45 | 0.033 | 14/52/08 | 68.3 | r. superior medial frontal gyrus |
|  |  |  |  | 26.4 | r. superior frontal gyrus |
|  |  |  |  | 4.0 | r. frontal pole |
|  |  |  |  | 1.3 | r. anterior cingulate gyrus |
|  | 940.05 | 0.033 | -02/50/02 | 30.5 | l. superior medial frontal gyrus |
|  |  |  |  | 27.8 | r. superior medial frontal gyrus |
|  |  |  |  | 26.2 | l. anterior cingulate gyrus |
|  |  |  |  | 8.2 | l. medial frontal cerebrum |
|  |  |  |  | 5.7 | r. anterior cingulate gyrus |
|  |  |  |  | 1.6 | r. medial frontal cerebrum |
| 3 | 862.88 | 0.049 | -02/36/22 | 50.5 | l. anterior cingulate gyrus |
|  |  |  |  | 21.0 | r. anterior cingulate gyrus |
|  |  |  |  | 16.0 | l. superior medial frontal gyrus |
|  |  |  |  | 12.4 | r. superior medial frontal gyrus |
| 9 | 861.79 | 0.049 | 33/26/51 | 89.0 | r. middle frontal gyrus |
|  |  |  |  | 8.1 | r. superior frontal gyrus |
|  |  |  |  | 2.9 | Background |

**Supplementary Table 4**

GMV: FWE-corrected clusters exhibiting significantly larger GMV in *nCSA* > *nCM*.

| *k* | TFCE | *p*FWE | coordinates (x/y/z) | % | anatomical region |
| --- | --- | --- | --- | --- | --- |
| 10619 | 1386.31 | 0.004 | -06/21/63 | 58.4 | l. superior frontal gyrus |
|  |  |  |  | 29.5 | l. cerebrum |
|  |  |  |  | 6.7 | r. cerebrum |
|  |  |  |  | 3.0 | background |
|  |  |  |  | 2.0 | l. superior medial frontal gyrus |
|  |  |  |  | 0.3 | r. superior frontal gyrus |
|  | 1320.98 | 0.005 | 04/16/26 | 49.9 | r. middle cingulate gyrus |
|  |  |  |  | 21.0 | l. middle cingulate gyrus |
|  |  |  |  | 14.0 | r. cerebral white matter |
|  |  |  |  | 10.8 | r. anterior cingulate gyrus |
|  |  |  |  | 1.6 | r. lateral ventricle |
|  |  |  |  | 1.6 | l. anterior cingulate gyrus |
|  |  |  |  | 1.1 | l. cerebral white matter |
|  | 1319.05 | 0.005 | 04/12/33 | 68.1 | r. middle cingulate gyrus |
|  |  |  |  | 22.5 | l. middle cingulate gyrus |
|  |  |  |  | 7.9 | r. cerebrum |
|  |  |  |  | 0.8 | r. cerebral white matter |
|  |  |  |  | 0.6 | l. cerebrum |
| 859 | 1067.21 | 0.018 | 32/-04/60 | 53.9 | r. precentral gyrus |
|  |  |  |  | 30.5 | r. middle frontal gyrus |
|  |  |  |  | 15.6 | r. superior frontal gyrus |
|  | 953.92 | 0.032 | 40/-02/44 | 83.3 | r. precentral gyrus |
|  |  |  |  | 16.7 | r. middle frontal gyrus |
|  | 922.17 | 0.037 | 28/-18/57 | 93.1 | r. precentral gyrus |
|  |  |  |  | 6.9 | r. postcentral gyrus |
| 989 | 1048.91 | 0.020 | -27/00/51 | 63.4 | l. middle frontal gyrus |
|  |  |  |  | 24.6 | l. superior frontal gyrus |
|  |  |  |  | 12.0 | l. precental gyrus |
|  | 1019.36 | 0.023 | -30/-04/62 | 45.7 | l. precentral gyrus |
|  |  |  |  | 33.9 | l. middle frontal gyrus |
|  |  |  |  | 20.5 | l. superior frontal gyrus |
|  | 990.35 | 0.027 | -20/-06/51 | 50.0 | l. superior frontal gyrus |
|  |  |  |  | 21.7 | l. cerebrum |
|  |  |  |  | 14.5 | l. precentral gyrus |
|  |  |  |  | 12.9 | l. middle frontal gyrus |
|  |  |  |  | 0.8 | l. cerebral white matter |
|  |  |  |  | 0.1 | l. middle cingulate gyrus |
| 1288 | 1041.91 | 0.021 | 30/38/-10 | 45.3 | r. anterior orbital gyrus |
|  |  |  |  | 22.2 | r. posterior orbital gyrus |
|  |  |  |  | 15.2 | r. medial orbital gyrus |
|  |  |  |  | 12.8 | r. lateral orbital gyrus |
|  |  |  |  | 3.3 | r. inferior frontal orbital gyrus |
|  |  |  |  | 0.6 | r. cerebral white matter |
|  |  |  |  | 0.4 | r. inferior frontal angular gyrus |
|  |  |  |  | 0.2 | r. anterior insula |
|  |  |  |  | 0.1 | r. middle frontal gyrus |
|  | 961.00 | 0.031 | 24/32/-26 | 40.6 | r. medial robital gyrus |
|  |  |  |  | 37.8 | Background |
|  |  |  |  | 20.7 | r. posterior orbital gyrus |
|  |  |  |  | 1.0 | r. temporal pole |
|  | 959.97 | 0.031 | 22/26/-18 | 50.4 | r. medial orbital gyrus |
|  |  |  |  | 45.1 | r. posterior orbital gyrus |
|  |  |  |  | 2.8 | r. anterior insula |
|  |  |  |  | 1.5 | r. cerebral white matter |
|  |  |  |  | 0.2 | r. gyrus rectus |
|  |  |  |  | 0.1 | Background |
| 781 | 957.70 | 0.031 | -48/20/00 | 41.5 | l. frontal operculum |
|  |  |  |  | 24.5 | l. inferior frontal angular gyrus |
|  |  |  |  | 23.2 | l. inferior frontal gyrus |
|  |  |  |  | 5.6 | l. inferior frontal orbital gyrus |
|  |  |  |  | 2.8 | l. anterior insula |
|  |  |  |  | 2.4 | l. temporal pole |
|  | 928.92 | 0.036 | -38/20/04 | 61.0 | l. frontal operculum |
|  |  |  |  | 28.1 | l. anterior insula |
|  |  |  |  | 7.8 | l. inferior frontal gyrus |
|  |  |  |  | 2.7 | l. inferior frontal orbital gyrus |
|  |  |  |  | 0.2 | l. inferior frontal angular gyrus |
|  | 920.29 | 0.037 | -39/16/14 | 43.8 | l. inferior frontal gyrus |
|  |  |  |  | 42.9 | l. frontal operculum |
|  |  |  |  | 4.9 | l. central operculum |
|  |  |  |  | 3.6 | l. inferior frontal angular gyrus |
|  |  |  |  | 2.5 | l. anterior insula |
|  |  |  |  | 2.2 | l. middle frontal gyrus |
| 47 | 898.98 | 0.041 | 39/21/32 | 89.9 | r. middle frontal gyrus |
|  |  |  |  | 10.1 | r. inferior frontal gyrus |
| 316 | 883.27 | 0.045 | 09/-46/26 | 66.4 | r. posterior cingulate gyrus |
|  |  |  |  | 26.8 | r. precuneus |
|  |  |  |  | 4.9 | r. cerebral white matter |
|  |  |  |  | 2.0 | l. posterior cingulate gyrus |
|  | 875.92 | 0.046 | 08/-45/14 | 59.4 | r. posterior cingulate gyrus |
|  |  |  |  | 25.1 | r. precuneus |
|  |  |  |  | 8.3 | r. cerebral white matter |
|  |  |  |  | 7.1 | l. posterior cingulate gyrus |
| 175 | 874.00 | 0.047 | 02/33/-15 | 35.2 | r. medial frontal cerebrum |
|  |  |  |  | 16.1 | l. medial frontal cerebrum |
|  |  |  |  | 15.2 | l. anterior cingulate gyrus |
|  |  |  |  | 15.0 | r. gyrus rectus |
|  |  |  |  | 12.0 | r. anterior cingulate gyrus |
|  |  |  |  | 4.9 | l. gyrus rectus |
|  |  |  |  | 1.0 | r. superior medial frontal gyrus |
|  |  |  |  | 0.7 | r. medial orbital gyrus |
|  | 874.00 | 0.047 | -04/27/-14 | 33.6 | l. anterior cingulate gyrus |
|  |  |  |  | 23.8 | l. medial frontal cerebrum |
|  |  |  |  | 17.1 | l. gyrus rectus |
|  |  |  |  | 12.3 | r. medial frontal cerebrum |
|  |  |  |  | 6.2 | r. anterior cingulate gyrus |
|  |  |  |  | 3.2 | l. subcallosal gyrus |
|  |  |  |  | 1.5 | l. medial orbital gyrus |
|  |  |  |  | 1.1 | l. cerebral white matter |
|  |  |  |  | 1.0 | r. gyrus rectus |
|  |  |  |  | 0.2 | r. subcallosal area |
| 33 | 871.83 | 0.047 | 26/-44/03 | 34.0 | r. lateral ventricle |
|  |  |  |  | 24.7 | r. cerebral white matter |
|  |  |  |  | 18.8 | r. lingual gyrus |
|  |  |  |  | 9.5 | r. posterior cingulate gyrus |
|  |  |  |  | 9.0 | r. hippocampus |
|  |  |  |  | 4.1 | r. precuneus |

**Supplementary Table 5**

GMV: FWE-corrected clusters exhibiting significantly larger GMV in *CSA* > *nCSA,* controlling for comorbidity (yes/no).

| *k* | TFCE | *p*FWE | coordinates (x/y/z) | % | anatomical region |
| --- | --- | --- | --- | --- | --- |
| 2443 | 930.61 | 0.012 | 33/-48/-30 | 87.9 | r. exterior cerebellum |
|  |  |  |  | 14.5 | r. cerebellum white matter |
|  |  |  |  | 6.6 | r. fusiform gyrus |
|  | 923.33 | 0.013 | 34/-56/-26 | 80.0 | r exterior cerebellum |
|  |  |  |  | 17.3 | r fusiform gyrus |
|  |  |  |  | 0.6 | r occipital fusiform gyrus |
|  |  |  |  | 0.2 | r cerebellum white matter |
|  | 901.00 | 0.015 | 51/-46/-30 | 44.5 | r inferior temporal gyrus |
|  |  |  |  | 43.7 | r exterior cerebellum |
|  |  |  |  | 8.1 | background |
|  |  |  |  | 3.8 | r fusiform gyrus |

**Supplementary Table 6**

GMV: FWE-corrected clusters exhibiting significantly larger GMV in *CSA* > *nCM,* controlling for comorbidity (yes/no).

| *k* | TFCE | *p*FWE | coordinates (x/y/z) | % | anatomical region |
| --- | --- | --- | --- | --- | --- |
| 217 | 1170.25 | 0.038 | -04/46/10 | 44.5 | l. anterior cingulate gyrus |
|  |  |  |  | 41.3 | l. superior medial frontal gyrus |
|  |  |  |  | 8.3 | r. superior medial frontal gyrus |
|  |  |  |  | 5.8 | r. anterior cingulate gyrus |
|  | 1106.37 | 0.048 | -02/50/02 | 30.5 | l. superior medial frontal gyrus |
|  |  |  |  | 27.8 | r. superior medial frontal gyrus |
|  |  |  |  | 26.2 | l. anterior cingulate gyrus |
|  |  |  |  | 8.2 | l. medial frontal cerebrum |
|  |  |  |  | 5.7 | r. anterior cingulate gyrus |
|  |  |  |  | 1.6 | r. medial frontal cerebrum |

**Supplementary Table 7**

GMV: FWE-corrected clusters exhibiting significantly larger GMV in *nCSA* > *nCM,* controlling for comorbidity (yes/no).

| *k* | TFCE | *p*FWE | coordinates (x/y/z) | % | anatomical region |
| --- | --- | --- | --- | --- | --- |
| 7741 | 1516.54 | 0.010 | -06/21/62 | 55.3 | l. superior frontal gyrus |
|  |  |  |  | 34.4 | l. cerebrum and motor |
|  |  |  |  | 7.0 | r. cerebrum and motor |
|  |  |  |  | 3.0 | l. superior medial frontal gyrus |
|  |  |  |  | 0.2 | Background |
|  |  |  |  | 0.1 | r. superior medial frontal gyrus |
|  | 1441.02 | 0.013 | 04/16/26 | 49.9 | r. middle cingulate gyrus |
|  |  |  |  | 21.0 | l. middle cingulate gyrus |
|  |  |  |  | 14.0 | r. cerebral white matter |
|  |  |  |  | 10.8 | r. anterior cingulate gyrus |
|  |  |  |  | 1.6 | r. lateral ventricle |
|  |  |  |  | 1.6 | l. anterior cingulate gyrus |
|  |  |  |  | 1.1 | l. cerebral white matter |
|  | 1439.20 | 0.013 | 04/12/33 | 68.1 | r. middle cingulate gyrus |
|  |  |  |  | 22.5 | l. middle cingulate gyrus |
|  |  |  |  | 7.9 | r. cerebrum and motor |
|  |  |  |  | 0.8 | r. cerebral white matter |
|  |  |  |  | 0.6 | l. cerebrum and motor |
| 1244 | 1344.70 | 0.018 | -06/-27/38 | 45.4 | l. middle cingulate gyrus |
|  |  |  |  | 40.0 | l. posterior cingulate gyrus |
|  |  |  |  | 4.9 | l. medial precentral gyrus |
|  |  |  |  | 4.0 | r. middle cingulate gyrus |
|  |  |  |  | 2.9 | r. posterior cingulate gyrus |
|  |  |  |  | 2.7 | l. cerebral white matter |
| 1135 | 1278.78 | 0.023 | -28/0/38 | 66.5 | l. middle frontal gyrus |
|  |  |  |  | 18.0 | l. superior frontal gyrus |
|  |  |  |  | 15.4 | l. precentral gyrus |
|  | 1251.06 | 0.026 | -32/-03/64 | 39.9 | l. precentral gyrus |
|  |  |  |  | 37.3 | l. middle frontal gyrus |
|  |  |  |  | 17.9 | l. superior frontal gyrus |
|  |  |  |  | 4.8 | Background |
|  | 1245.37 | 0.026 | -36/-02/57 | 54.8 | l. middle frontal gyrus |
|  |  |  |  | 45.1 | l. precentral gyrus |
|  |  |  |  | 0.1 | l. superior frontal gyrus |
| 443 | 1200.67 | 0.031 | 33/-04/60 | 59.1 | r. precentral gyrus |
|  |  |  |  | 31.0 | r. middle frontal gyrus |
|  |  |  |  | 9.8 | r. superior frontal gyrus |
|  | 1101.56 | 0.044 | 40/-02/44 | 83.3 | r. precentral gyrus |
|  |  |  |  | 16.7 | r. middle frontal gyrus |
| 731 | 1183.45 | 0.033 | 30/38/-09 | 44.5 | r. anterior orbital gyrus |
|  |  |  |  | 19.0 | r. posterior orbital gyrus |
|  |  |  |  | 12.1 | r. medial orbital gyrus |
|  |  |  |  | 11.8 | r. lateral orbital gyrus |
|  |  |  |  | 6.7 | r. inferior frontal orbital gyrus |
|  |  |  |  | 2.3 | r. cerebral white matter |
|  |  |  |  | 1.5 | r. inferior frontal angular gyrus |
|  |  |  |  | 1.4 | r. middle frontal gyrus |
|  |  |  |  | 0.6 | r. anterior insula |
|  | 1109.11 | 0.043 | 21/26/-21 | 59.7 | r. medial orbital gyrus |
|  |  |  |  | 35.7 | r. posterior orbital gyrus |
|  |  |  |  | 4.2 | Background |
|  |  |  |  | 0.3 | r. anterior insula |
|  |  |  |  | 0.1 | r. gyrus rectus |
|  | 1107.98 | 0.043 | 24/32/-26 | 40.6 | r. medial orbital gyrus |
|  |  |  |  | 37.8 | Background |
|  |  |  |  | 20.7 | r. posterior orbital gyrus |
|  |  |  |  | 1.0 | r. temporal pole |
| 543 | 1109.49 | 0.042 | 08/-46/18 | 55.9 | r. posterior cingulate gyrus |
|  |  |  |  | 31.6 | r. precuneus |
|  |  |  |  | 7.0 | l. posterior cingulate gyrus |
|  |  |  |  | 5.5 | r. cerebral white matter |
|  | 1109.40 | 0.042 | 09/-46/27 | 67.7 | r. posterior cingulate gyrus |
|  |  |  |  | 26.8 | r. precuneus |
|  |  |  |  | 3.6 | r. cerebral white matter |
|  |  |  |  | 2.0 | l. posterior cingulate gyrus |
|  | 1079.78 | 0.047 | 08/-56/26 | 76.6 | r. precuneus |
|  |  |  |  | 15.5 | r. posterior cingulate gyrus |
|  |  |  |  | 5.4 | l. precuneus |
|  |  |  |  | 1.5 | l. posterior cingulate gyrus |
|  |  |  |  | 0.8 | r. cuneus |
| 287 | 1105.45 | 0.043 | -48/20/0 | 41.5 | l. frontal operculum |
|  |  |  |  | 24.5 | l. inferior frontal angular gyrus |
|  |  |  |  | 23.2 | l. inferior frontal gyrus |
|  |  |  |  | 5.6 | l. inferior frontal orbital gyrus |
|  |  |  |  | 2.8 | l. anterior insula |
|  |  |  |  | 2.4 | l. temporal pole |
|  | 1087.96 | 0.046 | -39/21/03 | 59.8 | l. frontal operculum |
|  |  |  |  | 21.3 | l. anterior insula |
|  |  |  |  | 7.9 | l. inferior frontal gyrus |
|  |  |  |  | 7.0 | l. inferior frontal orbital gyrus |
|  |  |  |  | 4.0 | l. inferior frontal angular gyrus |
|  | 1079.78 | 0.047 | -36/21/15 | 46.8 | l. frontal operculum |
|  |  |  |  | 23.4 | l. inferior frontal gyrus |
|  |  |  |  | 16.7 | l. middle frontal gyrus |
|  |  |  |  | 9.8 | l. inferior frontal angular gyrus |
|  |  |  |  | 3.4 | l. anterior insula |
| 82 | 1085.35 | 0.046 | 36/52/-18 | 36.9 | Background |
|  |  |  |  | 36.9 | r. lateral orbital gyrus |
|  |  |  |  | 26.3 | r. anterior orbital gyrus |
| 82 | 1073.28 | 0.048 | 02/32/-15 | 34.2 | r. medial frontal cerebrum |
|  |  |  |  | 15.9 | l. medial frontal cerebrum |
|  |  |  |  | 15.6 | l. anterior cingulate gyrus |
|  |  |  |  | 15.3 | r. gyrus rectus |
|  |  |  |  | 12.9 | r. anterior cingulate gyrus |
|  |  |  |  | 4.9 | l. gyrus rectus |
|  |  |  |  | 0.8 | r. medial orbital gyrus |
|  |  |  |  | 0.5 | r. superior medial frontal gyrus |
| 23 | 1067.86 | 0.049 | 08/58/22 | 51.8 | r. superior medial frontal gyrus |
|  |  |  |  | 33.5 | r. superior frontal gyrus |
|  |  |  |  | 8.2 | r. frontal pole |
|  |  |  |  | 6.5 | l. superior medial frontal gyrus |

**Supplementary Table 8**

Cortical thickness: FWE-corrected clusters exhibiting significantly greater cortical thickness in *CSA* > *nCSA*.

| *k* | TFCE | *p*FWE | coordinates (x/y/z) | % | anatomical region |
| --- | --- | --- | --- | --- | --- |
| 11517 | 19182.83 | 0.011 | 31/20/08 | 9 | r. postcentral |
|  | 15282.46 | 0.020 | 49/08/13 | 9 | r. precentral |
|  | 14275.54 | 0.022 | 17/-96/-05 | 9 | r. superiorparietal |
|  |  |  |  | 9 | r. supramarginal |
|  |  |  |  | 8 | r. precuneus |
|  |  |  |  | 7 | r. insula |
|  |  |  |  | 7 | r. inferiorparietal |
|  |  |  |  | 7 | r. rostralmiddlefrontal |
|  |  |  |  | 6 | r. lateraloccipital |
|  |  |  |  | 5 | r. lingual |
|  |  |  |  | 4 | r. parsopercularis |
|  |  |  |  | 3 | r. parstriangularis |
|  |  |  |  | 2 | r. superiortemporal |
|  |  |  |  | 2 | r. cuneus |
|  |  |  |  | 2 | r. lateralorbitofrontal |
|  |  |  |  | 2 | r. inferiortemporal |
|  |  |  |  | 1 | r. isthmuscingulate |
|  |  |  |  | 1 | r. pericalcarine |
|  |  |  |  | 1 | r. parsorbitalis |
| 13137 | 16037.30 | 0.017 | -21/-83/39 | 12 | l. superiorfrontal |
|  | 15668.35 | 0.018 | -07/-74/11 | 9 | l. superiorparietal |
|  | 15324.55 | 0.019 | -21/-63/08 | 9 | l. precentral |
|  |  |  |  | 8 | l. supramarginal |
|  |  |  |  | 8 | l. inferiorparietal |
|  |  |  |  | 8 | l. precuneus |
|  |  |  |  | 7 | l. postcentral |
|  |  |  |  | 4 | l. insula |
|  |  |  |  | 4 | l. superiortemporal |
|  |  |  |  | 4 | l. caudalmiddlefrontal |
|  |  |  |  | 3 | l. pericalcarine |
|  |  |  |  | 3 | l. cuneus |
|  |  |  |  | 2 | l. lateraloccipital |
|  |  |  |  | 2 | l. posteriorcingulate |
|  |  |  |  | 2 | l. lingual |
|  |  |  |  | 2 | l. rostralanteriorcingulate |
|  |  |  |  | 2 | l. caudalanteriorcingulate |
|  |  |  |  | 2 | l. parsopercularis |
|  |  |  |  | 2 | l. lateralorbitofrontal |
|  |  |  |  | 2 | l. rostralmiddlefrontal |
|  |  |  |  | 1 | l. medialorbitofrontal |
|  |  |  |  | 1 | l. parstriangularis |
|  |  |  |  | 1 | l. parsorbitularis |
|  |  |  |  | 1 | l. bankssts |
| 246 | 9500.77 | 0.049 | 06/44/26 | 100 | r. superiorfrontal |

**Supplementary Table 9**

Cortical thickness: FWE-corrected clusters exhibiting significantly greater cortical thickness in *CSA* > *nCM*.

| *k* | TFCE | *p*FWE | coordinates (x/yz/) | % | anatomical region |
| --- | --- | --- | --- | --- | --- |
| 13400 | 27566.17 | 0.004 | -29/04/54 | 15 | l. superiorfrontal |
|  | 27506.49 | 0.004 | -48/10/-01 | 9 | l. supramarginal |
|  | 26771.16 | 0.004 | -42/01/39 | 8 | l. precuneus |
|  |  |  |  | 7 | l. superiortemporal |
|  |  |  |  | 6 | l. inferiorparietal |
|  |  |  |  | 6 | l. superorparietal |
|  |  |  |  | 5 | l. caudalmiddlefrontal |
|  |  |  |  | 5 | l. insula |
|  |  |  |  | 4 | l. rostralmiddlefrontal |
|  |  |  |  | 4 | l. precentral |
|  |  |  |  | 4 | l. parsopercularis |
|  |  |  |  | 3 | l. postcentral |
|  |  |  |  | 3 | l. parstriangularis |
|  |  |  |  | 3 | l. lateraloccipital |
|  |  |  |  | 2 | l. pericalcerine |
|  |  |  |  | 2 | l. cuneus |
|  |  |  |  | 2 | l. medialorbitofrontal |
|  |  |  |  | 2 | l. inferiortemporal |
|  |  |  |  | 1 | l. transversetemporal |
|  |  |  |  | 1 | l. posteriorcingulate |
|  |  |  |  | 1 | l. rostralanteriorcingulate |
|  |  |  |  | 1 | l. lateralorbitofrontal |
| 6478 | 18517.88 | 0.018 | 63/-40/24 | 16 | r. supramarginal |
|  | 18504.70 | 0.018 | 53/-49/26 | 15 | r. superiortemporal |
|  | 18393.23 | 0.018 | 33/10/10 | 11 | r. inferiorparietal |
|  |  |  |  | 10 | r. insula |
|  |  |  |  | 9 | r. rostralmiddlefrontal |
|  |  |  |  | 8 | r. parsopercularis |
|  |  |  |  | 7 | r. parstriangularis |
|  |  |  |  | 6 | r. lateralorbitofrontal |
|  |  |  |  | 5 | r. precentral |
|  |  |  |  | 3 | r. caudalmiddlefrontal |
|  |  |  |  | 3 | r. middletemporal |
|  |  |  |  | 2 | r. parsorbitalis |
|  |  |  |  | 2 | r. inferiortemporal |
|  |  |  |  | 2 | r. postcentral |
|  |  |  |  | 2 | r. transversetemporal |
|  |  |  |  | 2 | bankssts |

**Supplementary Table 10**

Cortical thickness: FWE-corrected clusters exhibiting significantly greater cortical thickness in *CSA* > *nCSA.* controlling for symptom severity (HAMD sum score).

| *k* | TFCE | *p*FWE | coordinates (x/y/z) | % | anatomical region |
| --- | --- | --- | --- | --- | --- |
| 1145 | 20488.59 | 0.027 | 31/20/08 | 59 | r. insula |
|  |  |  |  | 13 | r. lateraloribtofrontal |
|  |  |  |  | 10 | r. parstriangularis |
|  |  |  |  | 7 | r. supramarginal |
|  |  |  |  | 7 | r. parsopercularis |
|  |  |  |  | 7 | r. parsorbitalis |
| 372 | 17029.53 | 0.047 | 55/-05/39 | 80 | r. precentral |
|  | 16914.12 | 0.048 | 49/08/13 | 20 | r. parsopercularisl |
| 844 | 18089.93 | 0.040 | -29/-69/25 | 42 | l. superioroparietal |
|  |  |  |  | 20 | l. cuneus |
|  |  |  |  | 18 | l. precuneus |
|  |  |  |  | 15 | l. inferiorparietal |
|  |  |  |  | 6 | l. pericalcarine |

**Supplementary Table 11**

Cortical thickness: FWE-corrected clusters exhibiting significantly greater cortical thickness in *CSA* > *nCM.* controlling for symptom severity (HAMD sum score).

| *k* | TFCE | *p*FWE | coordinates (x/y/z) | % | anatomical region |
| --- | --- | --- | --- | --- | --- |
| 20782 | 29376.29 | 0.000 | -46/37/-02 | 11 | l. superiorfrontal |
|  | 28901.12 | 0.000 | -29/04/54 | 7 | l. supramarginal |
|  | 27970.67 | 0.000 | -63/-16/19 | 7 | l. postcentral |
|  | 27947.76 | 0.000 | -58/-27/17 | 6 | l. superiorparietal |
|  | 27920.00 | 0.000 | -42/01/39 | 6 | l. superiortemporal |
|  | 27109.10 | 0.000 | -61/-08/01 | 6 | l. precuneus |
|  | 26471.05 | 0.000 | -54/-55/23 | 6 | l. inferiorparietal |
|  | 22611.52 | 0.000 | -29/33/38 | 5 | l. rostralmiddlefrontal |
|  | 22114.99 | 0.000 | -44/-75/32 | 5 | l. precentral |
|  | 18547.31 | 0.000 | -51/-20/-13 | 4 | l. lateraloccipital |
|  | 18424.96 | 0.000 | -07/-79/11 | 4 | l. insula |
|  | 17986.80 | 0.001 | -22/-81/35 | 4 | l. caudalmiddlefrontal |
|  | 17870.29 | 0.001 | -06/-52/46 | 3 | l. inferiorfrontal |
|  | 17866.77 | 0.001 | -37/42/15 | 3 | l. parsopercularis |
|  | 17542.25 | 0.001 | -16/-101/11 | 3 | l. middletemporal |
|  | 17640.80 | 0.001 | -51/-15/-39 | 2 | l. parscentral |
|  |  |  |  | 2 | l. parstriangularis |
|  |  |  |  | 2 | l. fusiform |
|  |  |  |  | 2 | l. pericalcarine |
|  |  |  |  | 2 | l. cuneus |
|  |  |  |  | 1 | l. posteriorcingulate |
|  |  |  |  | 1 | l. medialorbitofrontal |
|  |  |  |  | 1 | bankssts |
|  |  |  |  | 1 | l. lateralorbitofrontal |
| 18647 | 19948.98 | 0.000 | 64/-40/23 | 9 | r. superiorfrontal |
|  | 19237.85 | 0.000 | 47/-56/34 | 8 | r. supramarginal |
|  | 18980.77 | 0.000 | 33/10/10 | 7 | r. inferiorparietal |
|  | 16702.84 | 0.000 | 42/28/19 | 7 | r. rostralmiddlefrontal |
|  | 15520.37 | 0.000 | 39/50/15 | 6 | r. superiortemporal |
|  | 13187.93 | 0.001 | 50/-38/45 | 6 | r. postcentral |
|  | 12709.39 | 0.001 | 50/-53/14 | 5 | r. precuneus |
|  | 11774.00 | 0.001 | 41/-40/37 | 5 | r. precentral |
|  |  |  |  | 5 | r. superiorparietal |
|  |  |  |  | 4 | r. insula |
|  |  |  |  | 4 | r. lateraloccipital |
|  |  |  |  | 3 | r. middletemporal |
|  |  |  |  | 3 | r. paracentral |
|  |  |  |  | 3 | r. lingual |
|  |  |  |  | 3 | r. parsopercularis |
|  |  |  |  | 3 | r. lateralorbitofrontal |
|  |  |  |  | 3 | r. parstriangularis |
|  |  |  |  | 3 | r. inferiortemporal |
|  |  |  |  | 3 | r. caudalmiddlefrontal |
|  |  |  |  | 2 | r. pericalcarine |
|  |  |  |  | 2 | r. cuneus |
|  |  |  |  | 1 | bankssts |
|  |  |  |  | 1 | r.parsorbitalis |

**Supplementary Table 12**

Cortical thickness: FWE-corrected clusters exhibiting significantly greater cortical thickness in *CSA* > *nCSA.* controlling for comorbidity (yes/no).

| *k* | TFCE | *p*FWE | coordinates (x/y/z) | % | anatomical region |
| --- | --- | --- | --- | --- | --- |
| 13467 | 19298.19 | 0.007 | 31/20/08 | 9 | r. precentral |
|  | 15722.28 | 0.013 | 57/-03/36 | 8 | r. superiorfrontal |
|  | 14241.98 | 0.017 | 17/-96/-05 | 8 | r. postcentral |
|  |  |  |  | 8 | r. supramarginal |
|  |  |  |  | 8 | r. superiorparietal |
|  |  |  |  | 7 | r. precuneus |
|  |  |  |  | 6 | r. insula |
|  |  |  |  | 6 | r. inferiorparietal |
|  |  |  |  | 6 | r. rostralmiddlefrontal |
|  |  |  |  | 6 | r. lateraloccipital |
|  |  |  |  | 4 | r. lingual |
|  |  |  |  | 3 | r. parsopercularis |
|  |  |  |  | 3 | r. fusiform |
| 14272 | 16102.46 | 0.012 | -05/-78/24 | 11 | l. superiorfrontal |
|  | 15501.70 | 0.014 | -05/-64/37 | 9 | l. precentral |
|  | 14412.51 | 0.016 | -08/-40/45 | 9 | l. superiorparietal |
|  |  |  |  | 8 | l. supramarginal |
|  |  |  |  | 8 | l. inferiorparietal |
|  |  |  |  | 7 | l. precuneus |
|  |  |  |  | 7 | l. postcentral |
|  |  |  |  | 4 | l. superiormatginal |
|  |  |  |  | 4 | l. insula |
|  |  |  |  | 4 | l. caudalmiddelfontal |
|  |  |  |  | 3 | l. pericalcarine |
|  |  |  |  | 2 | l. lateraloccipital |
|  |  |  |  | 2 | l. cuneus |
|  |  |  |  | 2 | l. posteriorcingulate |
|  |  |  |  | 2 | l. rostralmiddlefrontal |
|  |  |  |  | 2 | l. lingual |
|  |  |  |  | 2 | l. lateralorbitofrontal |
|  |  |  |  | 2 | l. parsopercularis |
|  |  |  |  | 2 | l. caudalanteriorcingulate |
|  |  |  |  | 2 | l. rostralorbitofrontal |
|  |  |  |  | 1 | l. medialorbitofrontal |
|  |  |  |  | 1 | l. parstriangularis |
|  |  |  |  | 1 | l. parsobitalis |
|  |  |  |  | 1 | bankssts |

**Supplementary Table 13**

Cortical thickness: FWE-corrected clusters exhibiting significantly greater cortical thickness in *CSA* > *nCM.* controlling for comorbidity (yes/no).

| *k* | TFCE | *p*FWE | coordinates (x/y/z) | % | anatomical region |
| --- | --- | --- | --- | --- | --- |
| 3086 | 27000.48 | 0.038 | -48/10/-01 | 18 | l. caudalmiddlefrontal |
|  | 25875.31 | 0.041 | -39/06/56 | 17 | l. insula |
|  | 25436.81 | 0.043 | -59/-09/-06 | 12 | l. parsopercularis |
|  |  |  |  | 9 | l. precentral |
|  |  |  |  | 9 | l. superiortemporal |
|  |  |  |  | 9 | l. parstriangularis |
|  |  |  |  | 8 | l. supramarginal |
|  |  |  |  | 7 | l. postcentral |
|  |  |  |  | 6 | l. transversetemporal |
|  |  |  |  | 3 | l. rostralmiddlefrontal |
|  |  |  |  | 1 | l. superiorfrontal |
| 316 | 24699.18 | 0.045 | -51/-54/41 | 89 | l. supramarginal |
|  | 24048.91 | 0.048 | -54/-55/23 | 11 | l. inferiorparietal |
